# A six-plex digital PCR assay for monitoring respiratory viruses in wastewater

**DOI:** 10.1101/2024.12.06.24317241

**Authors:** Melissa Pitton, Rachel E. McLeod, Lea Caduff, Ayazhan Dauletova, Jolinda de Korne-Elenbaas, Charles Gan, Camille Hablützel, Aurélie Holschneider, Seju Kang, Guy Loustalot, Patrick Schmidhalter, Linda Schneider, Anna Wettlauffer, Daniela Yordanova, Timothy R. Julian, Christoph Ort

**Author notes:** Authors contributed equally (Melissa Pitton, Rachel E. McLeod). Authors contributed equally (Timothy R. Julian, Christoph Ort). Corresponding authors: Christoph Ort, Timothy R. Julian.

## Abstract

Wastewater-based surveillance systems can track trends in multiple pathogens simultaneously by leveraging efficient, streamlined laboratory processing. In Switzerland, wastewater surveillance is conducted for fourteen locations representing 2.3 million people, or 26% of the national population, with simultaneous surveillance of four respiratory pathogens. Trends in respiratory diseases are tracked using a novel, six-plex digital PCR assay targeting Influenza A, Influenza B, Respiratory Syncytial Virus, and SARS-CoV-2 N1 and N2 genes, as well as Murine Hepatitis Virus for recovery efficiency control. The multiplex assay was developed to ensure sensitivity and accurate quantification for all targets simultaneously. Wastewater data is also integrated with disease data obtained through both a mandatory disease reporting system and the Swiss Sentinel System (Sentinella), a voluntary reporting system for general practitioners. Comparisons between wastewater data and case data from July 2023 through July 2024 demonstrate a high level of agreement, specifically for Influenza A, SARS-CoV-2, and Respiratory Syncytial Virus. Lower correspondence is observed for Influenza B, which highlights challenges in tracking disease dynamics during seasons without pronounced outbreak periods. Wastewater monitoring further revealed that targeting the N1 or N2 gene led to divergent estimates of SARS-Cov-2 viral loads, highlighting the impact of mutations in the target region of the assay on tracking trends. The study emphasizes the importance of an integrated wastewater monitoring program as a complementary tool for public health surveillance by demonstrating clear concordance with clinical data for respiratory pathogens beyond SARS-CoV-2.

## INTRODUCTION

Respiratory infections are a leading cause of morbidity and mortality worldwide – particularly in children and the elderly^1^. Such infections are commonly caused by viral pathogens, including Influenza A (IAV) and B (IBV), Respiratory Syncytial Virus (RSV), and SARS-CoV-2. Infections from respiratory viruses typically follow a seasonal pattern, with one or two peaks annually, though in many countries influenza transmission is identified year-round^2^. Our knowledge about respiratory virus epidemiology and infection dynamics is overwhelmingly driven by clinical data reporting systems. However, such systems can be subject to bias, likely underestimating the disease prevalence^3^, or incompletely capturing dynamics due to changes in testing or reporting strategies or changes in population behavior seeking medical care.

Wastewater-based surveillance (WBS) is proposed as an alternative and/or complementary option for tracking respiratory viruses^4,5^. WBS relies on wastewater analysis for detecting genetic material of a target pathogen, often by implementing molecular-based assays, such as quantitative PCR (qPCR) and digital PCR (dPCR). The COVID-19 pandemic has highlighted the potential and advantages of WBS^6,7^, which is now applied worldwide.

WBS allows a broad, population-level perspective by sampling wastewater, offering insights into the trends of entire communities or regions from just one 24-hour composite wastewater sample per day. Further, WBS is useful as a complementary tool to traditional surveillance methods, providing data that can enhance the accuracy and timeliness of public health interventions^8–10^. Indeed, previous studies reported a strong association between wastewater viral concentrations and reported clinical cases, suggesting that when both surveillance strategies are sensitive enough, the resulting data solidly correspond.

WBS has been applied to the detection of diverse respiratory pathogens beyond SARS-CoV-2, including RSV and influenza viruses^11–13^. Tracking pathogen concentrations in settled solids and wastewater influent aligns with available clinical data, which often relies on metrics such as clinical positive rates (as in Hughes et al. for RSV) or influenza-like illnesses (as in Wolfe et al. and Zheng et al. for Influenza A). Moreover, simultaneous detection of multiple pathogens in wastewater offers an efficient means of monitoring distinct diseases. To achieve this multiplexing of pathogens, dPCR, compared to qPCR, offers substantial potential^14^.

The wastewater monitoring program in Switzerland was initiated in July 2020 for the surveillance of SARS-CoV-2, and subsequently expanded for the detection of IAV, IBV, and RSV, with the implementation of a second multiplex dPCR assay^15^ in November 2022. To improve the workflow and reduce costs of the surveillance program, we developed a novel multiplex dPCR assay for detecting simultaneously the four target respiratory viral pathogens (IAV, IBV, RSV, and SARS-CoV-2) as well as an additional quality control to measure recovery efficiency (Murine Hepatitis Virus, MHV). Here, we report the development and application of the multiplex dPCR assay to the established wastewater surveillance framework for monitoring in Switzerland.

## RESULTS

### Performance of the six-plex dPCR assay and quality control indicators

The six-plex dPCR assay, referred to as RESPV6, was designed for detecting six target genes with specific fluorophores: SARS-CoV-2 N1 gene (SARS-N1, ATTO425), SARS-CoV-2 N2 gene (SARS-N2, FAM), RSV N gene (RSV-N, ROX), IAV M gene (IAV-M, Cy5), IBV M gene (IBV-M, HEX), and MHV M gene (MHV-M, Cy5.5). All selected targets produced clusters which were separable, allowing for discrimination between positive and negative partitions, and between distinct targets (**Fig. 1**).

**Figure 1.**
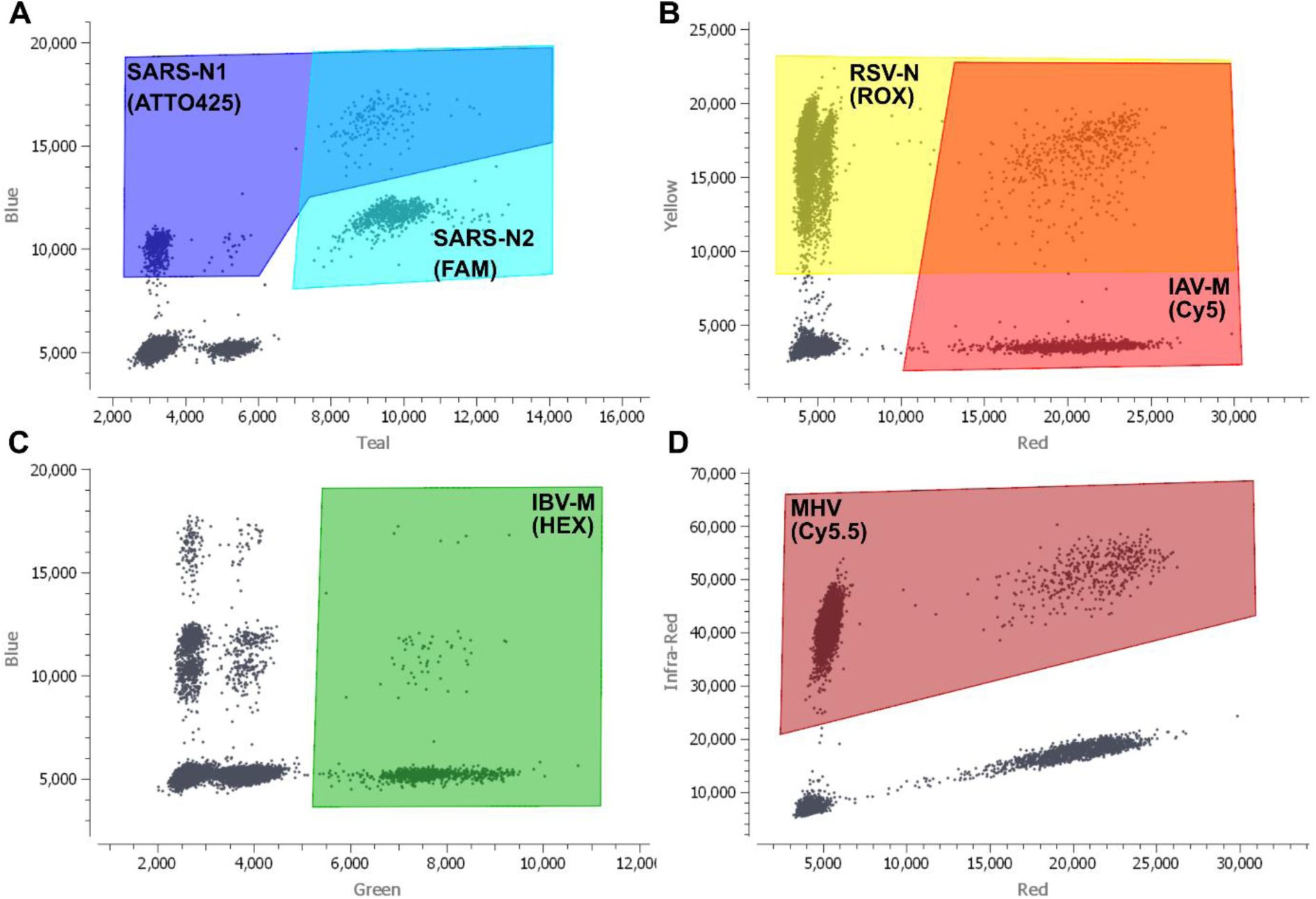
Two-dimensional visualization of the partition classification of RESPV6 dPCR assay after measuring positive control. Both x-axes and y-axes show fluorescence intensities. Titles on axes indicate the fluorescent channel. Each black dot corresponds to a dPCR partition. Partitions are classified by manually drawing polygons. Investigated targets are shown in bold within polygons with fluorophores in parentheses. Partitions enclosed in the area where polygons overlap are positive for multiple targets (i.e., one partition contains two or more targets). **(A)** SARS-CoV-2 genes N1 and N2 can both be gated in teal and blue channels. **(B)** RSV-N and IAV-M can be gated in yellow and red channels, respectively. **(C)** IBV-M positive clusters are separable in the green channel. **(D)** MHV is gated in the infra-red channel. This figure has been formatted using Inkscape.

The assay was applied for monitoring wastewater between July 2023 and July 2024, leading to a total of 13’028 individual dPCR reactions. Of those reactions, 7’795 were performed on wastewater samples collected from 14 WWTPs (**Supplemental Table 1**), 2’482 were run for control purposes, and 2’751 for assessing PCR inhibition. Altogether, we analyzed 3’538 distinct wastewater samples.

Since wastewater is a complex matrix and can contain inhibitors we determined the levels of PCR inhibition present in the analyzed wastewater nucleic acid extracts, and repeated samples showing more than 40% inhibition. For reactions that passed quality control (7’052/7’795), we observed a median PCR inhibition of 3% (interquartile range [IQR] 14%). We observed a significant variation across different locations (Kruskal-Wallis test, *p* < 0.001). The Dunn’s multiple comparisons test displayed that samples from Basel had a significantly higher level of inhibition (median of 7%) than other WWTPs, whereas samples from Solothurn had lower level of inhibition compared to other WWTPs (median of −1%) (**Supplemental Table S2**).

During this study, we observed dPCR reaction failure in a total of 743 reactions out of 7’795 (10%). Of these, 121 failed because of high levels of inhibition (>40% inhibition, 16%), 303 due to a failed no template control suggesting possible contamination of the sample (NTC, 40%), and 334 for insufficient number of partitions (i.e., <15’000 droplets, 44%). Reaction failure resulted in data for 281 samples (8%) being taken from a single replicate rather than the average of duplicates.

Based on the recovery of MHV, the recovery efficiency control, we observed a median recovery rate of 18% (IQR 38%) and an average of 30% (± 27%), which was derived from running efficiency control in 3’951 dPCR reactions. We detected a significant difference among locations (Kruskal-Wallis test, *p* < 0.001). The Dunn’s multiple comparisons test showed that samples from Basel had a significantly lower extraction efficiency than other WWTPs (**Supplemental Table S3**).

Among the reactions containing wastewater extracts that passed quality control, we observed a median number of partitions of 22’104 (IQR 4’726). Moreover, we estimated the volume of reaction mixture that did not form partitions (dead volume) which showed that the median percentage of dead volume for each reaction was 54% (IQR 10%).

### Detection of viral pathogens in Swiss wastewater

Our dPCR analysis of 3’538 samples revealed that IAV was detected in 27% of the samples (n = 941), IBV in 42% (n = 1’491), and RSV in 38% of the samples (n = 1’338). SARS-CoV-2 was detected in 99% and 98% of the samples for the N1 and N2 genes respectively (n = 3’499, n = 3’472). Despite N1 and N2 displaying a marked positive linear relationship (R^2^ = 0.97), N2 values were generally higher than N1 values (**Fig. 2**). Indeed, the ratio of N1 to N2 was 0.87 ± 0.33 (mean ± standard deviation [s.d.]) (n = 3’538). Interestingly, we noted a shift in the ratio of N1 to N2 over the monitoring period. From July 2023 to January 2024 the ratio was constantly below one at all locations (0.71 ± 0.23, n = 1’648), but then returned to one after January 2024 (1.01 ± 0.35, n = 1’890) (**Supplemental Fig. S1**).

**Figure 2.**
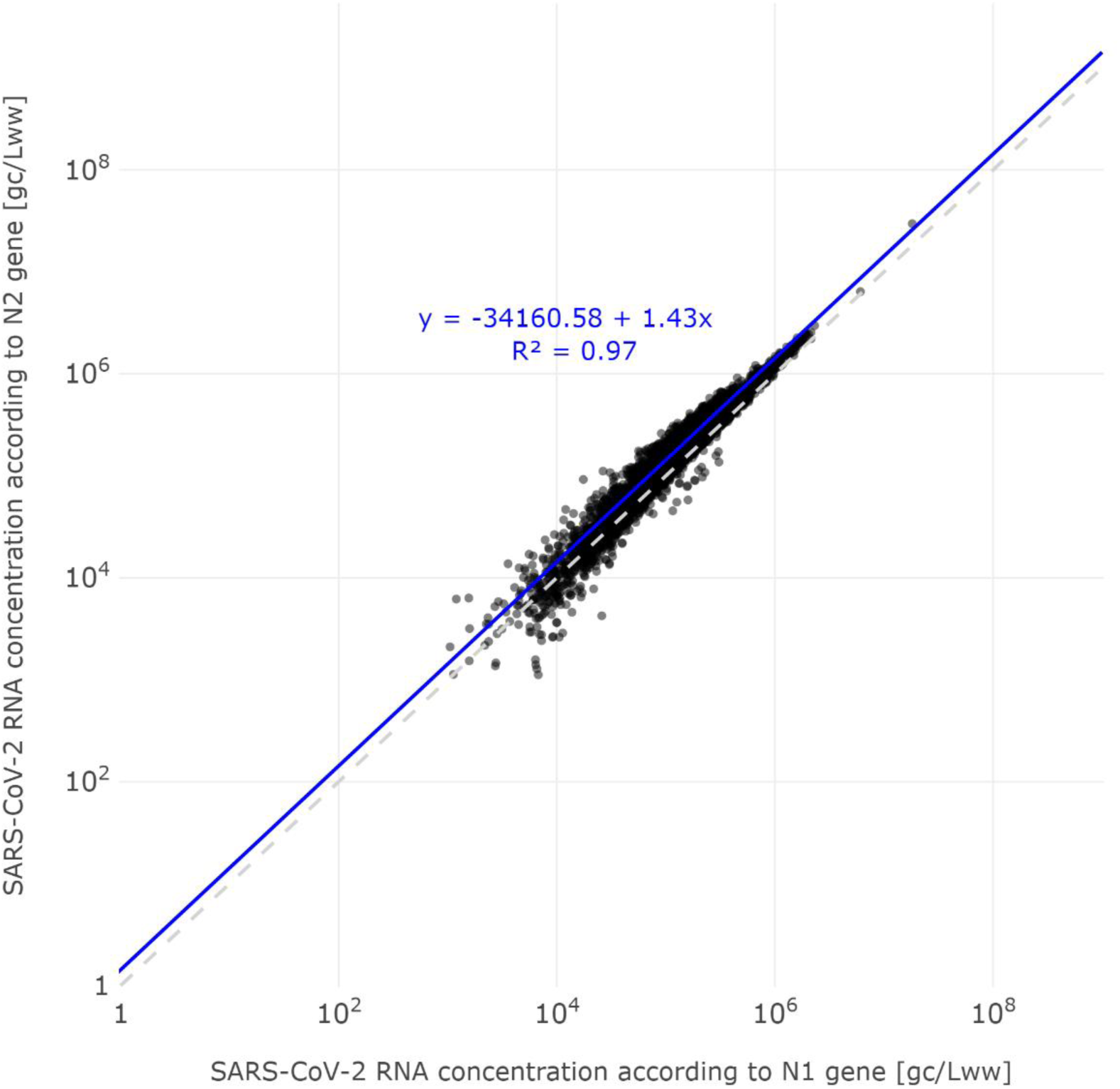
SARS-CoV-2 RNA concentrations comparing N1 and N2 genes. Concentrations are expressed in gene copies per liter of wastewater (gc/Lww) and shown on a log scale. The grey dashed line indicates the identity line. The linear regression line is shown in blue.

### Viral loads of respiratory pathogens in Swiss wastewater

We explored the epidemiology of the target respiratory viruses by observing the viral loads in wastewater, expressed in gene copies per person per day (gc person^-1^ day^-1^), over time. To better identify and interpret the trends, we calculated the median value over a centrally aligned seven-day rolling period. This helped smooth fluctuations and outliers.

This observational period was strongly characterized by a considerable wave of SARS-CoV-2 loads, detected in all 14 catchments, from October 2023 until February 2024 (**Fig. 3**). Each location was characterized by an outbreak peak (i.e., maximum median value) between 9^th^ November 2023 and 21^st^ December 2023. Lucerne displayed the highest load of 9.7 × 10^8^ gc person^-1^ day^-1^ on the 8^th^ of December 2023 (**Fig. 3C**). Importantly, a second peak of SARS-CoV-2 was identified in all catchments starting in April 2024, particularly in Lausanne, where a clear second wave was visible and comparable to the previous one (**Fig. 3N**).

**Figure 3.**
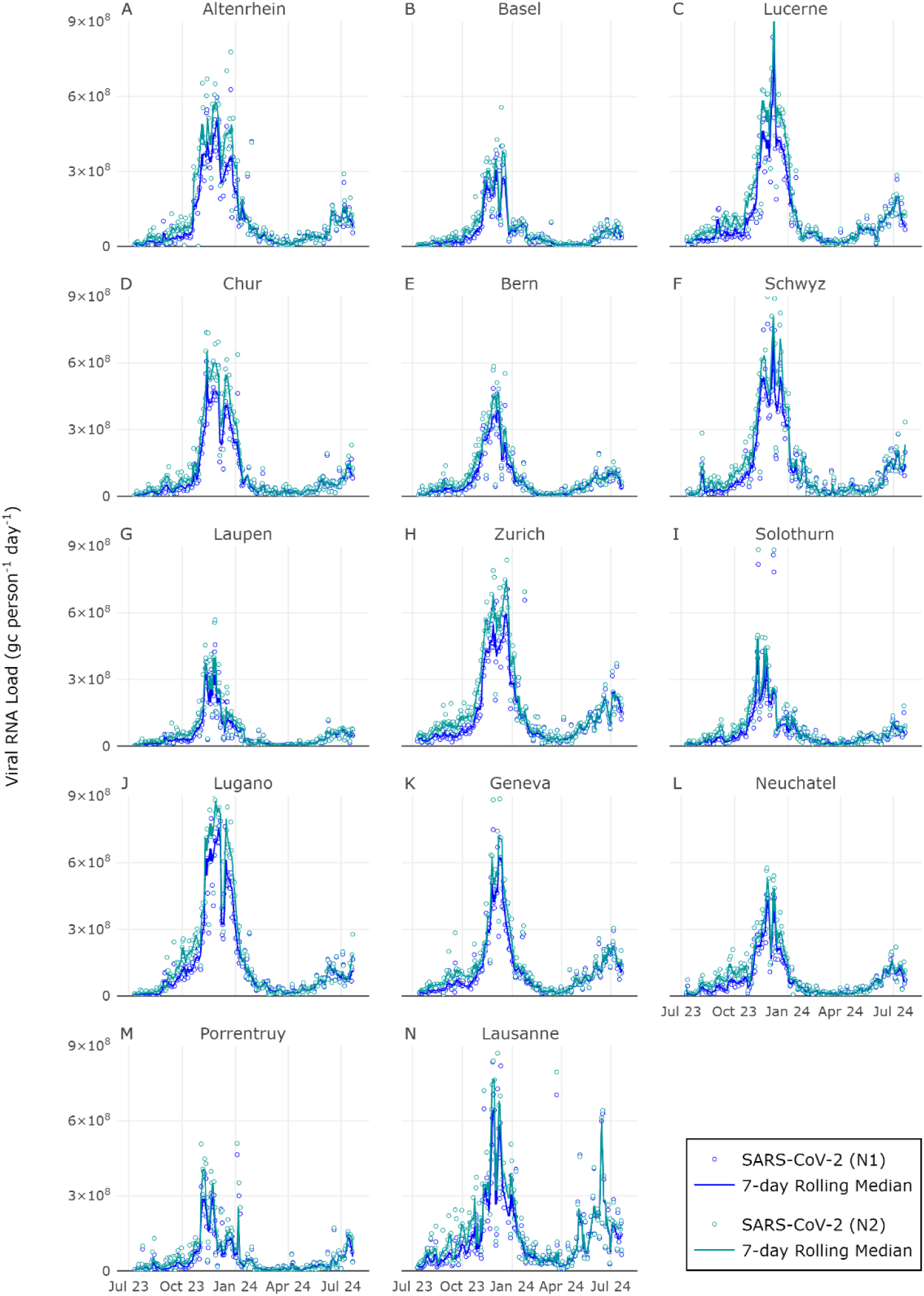
SARS-CoV-2 N1 and N2 load in wastewater across 14 Swiss locations. SARS-CoV-2 RNA loads, expressed in gene copies (gc) per person per day, were calculated for N1 and N2 genes. Measured RNA loads are represented with a circle (blue for SARS-N1 and teal for SARS-N2), whereas the respective seven-day rolling medians are displayed with solid lines. Each panel indicates a different WWTP. For visualization purposes, some very high load values are not visible in the plots and are listed in **Supplemental Table S7**.

The surveillance period was also characterized by the presence of the other measured pathogens IAV, IBV, and RSV, though the concentrations and loads in wastewater were considerably lower than those of SARS-CoV-2, differing by more than one order of magnitude (**Fig. 3**, **Fig. 4**). Between December 2023 and mid-March 2024, we encountered a seasonal outbreak of IAV. Maximum peaks were seen within a one-month period (18^th^ Jan – 16^th^ Feb 2024) for 13 locations. The exception was Lugano where the peak occurred earlier (**Supplemental Fig. S2A**). Moreover, Lugano had the highest peak value for IAV among all locations, with 5.1 × 10^7^ gc person^-1^ day^-1^ on the 28^th^ of December 2023 (**Fig. 4J**). Between November 2023 and May 2024, we observed a wave of RSV, and peak values were identified between 3^rd^ December 2023 and 15^th^ February 2024 for 13 locations, except for Solothurn which was characterized by a peak in July 2023 (**Fig. 4I** and **Fig. S2C**).

**Figure 4.**
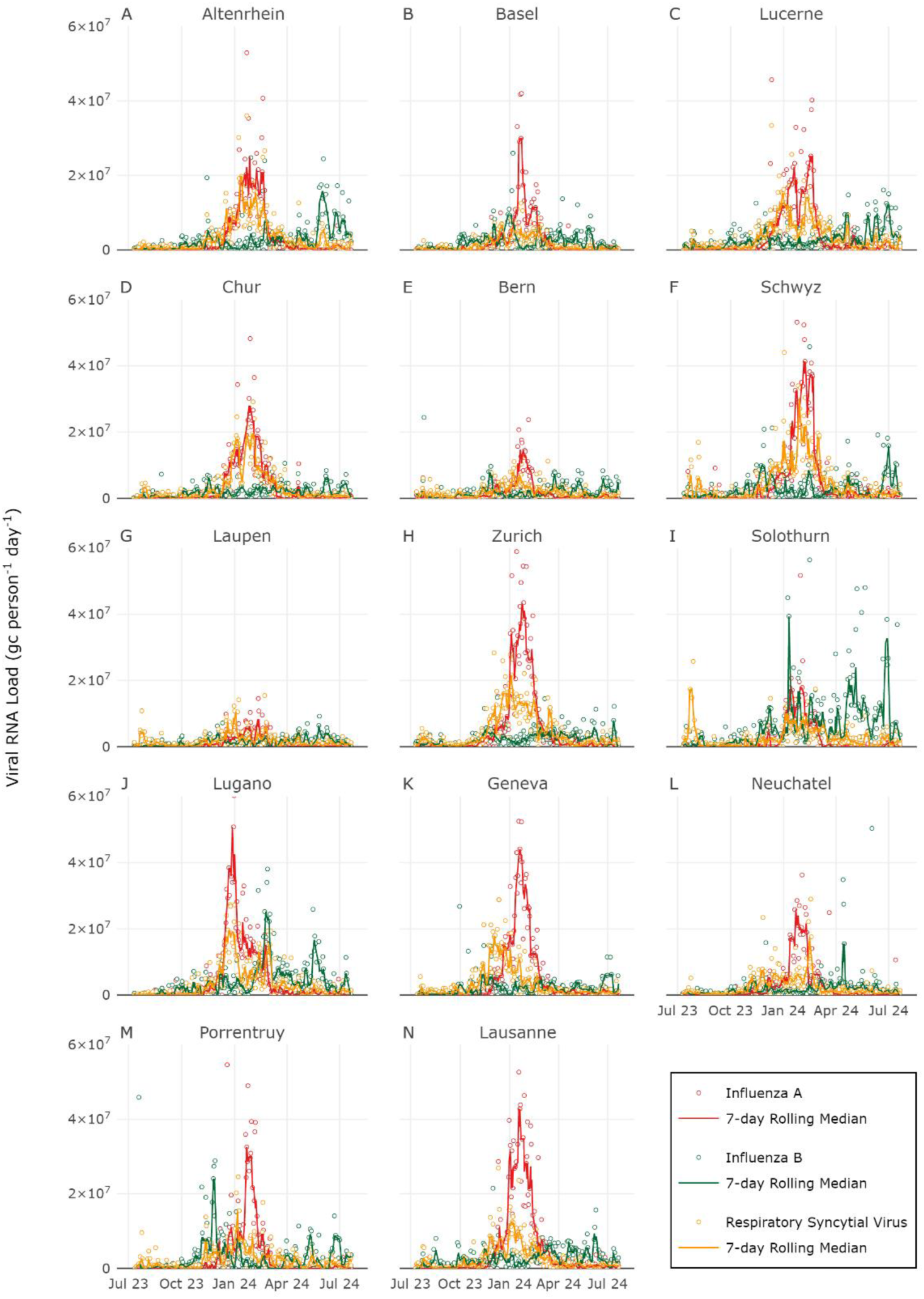
Influenza A, Influenza B and RSV RNA loads in wastewater across 14 Swiss locations. Viral RNA loads were expressed in gene copies (gc) per person per day. Measured RNA loads are represented with a circle (red for IAV, green for IBV, and yellow for RSV), whereas the respective seven-day rolling median is displayed with solid lines. Each panel indicates a different WWTP. For visualization purposes, some very high load values are not visible in the plots and are listed in **Supplemental Table S7**.

Unlike SARS-CoV-2, IAV, and RSV outbreaks, which showed similar and comparable trends across different Swiss locations, the trends for IBV were less clear, with fewer comparable and distinguishable patterns among catchments (**Supplemental Fig. S2B**). As such we observe the peak values for IBV over more than eight months (13^th^ Nov 2023 – 12^th^ Jul 2024).

### Comparison of viral loads in wastewater with clinical data from different sources

At the national level, we observed consistent relationships between weekly wastewater viral loads and weekly clinical cases from the Sentinella system for all four pathogens tracked (**Fig. 5**). We observed a strong positive correlation between IAV RNA loads in wastewater and IAV clinical cases (Pearson correlation, r = 0.95 [*p* < 0.0001]), between SARS-CoV-2 RNA loads and clinical cases (r = 0.87 [*p* < 0.0001]), and between RSV RNA loads and clinical cases (r = 0.68 [*p* < 0.0001]). The same analysis for IBV showed a rather low correlation (r = 0.28 [*p* = 0.04]) (**Fig. 5B**).

**Figure 5.**
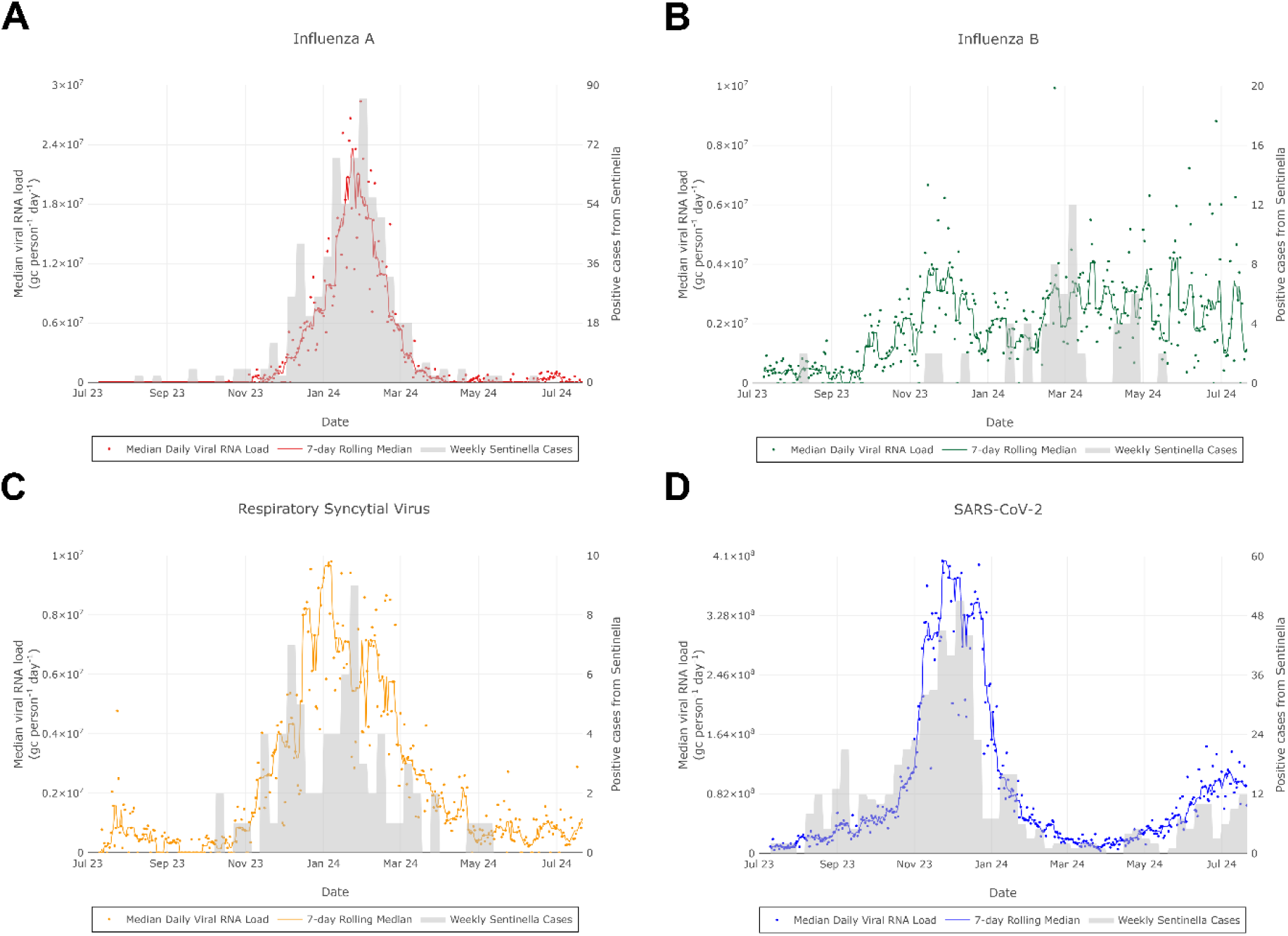
Comparison of viral RNA loads in wastewater and number of cases from Sentinella system. Positive cases, expressed in absolute numbers, refer to the entire Swiss territory, and were taken from the Sentinella database. Cases are shown using light grey bars. Some high load values are beyond the plotted y-max values. They are listed in **Supplemental Table S7**. This figure has been formatted using Inkscape.

Subsequently, we performed the same correlation coefficient analysis using clinical data publicly available from the mandatory reporting system, which reports clinical positive cases per week for IAV, IBV, and SARS-CoV-2. As expected, we observed higher correlation coefficients (r = 0.99 for IAV [*p* < 0.0001], r = 0.44 for IBV [*p* = 0.0008], and r = 0.93 for SARS-CoV-2 [*p* < 0.0001]), likely due to the better spatial resolution of such data which is less prone to bias (**Supplemental Fig. S3**). Additionally, we investigated the relationship between clinical data from Sentinella and from the mandatory reporting system, and we observed very strong Pearson correlation coefficients for SARS-CoV-2 and IAV (r = 0.92 [*p* < 0.0001] and r = 0.95 [*p* < 0.0001]). However, the correlation for IBV was only moderate (r = 0.63 [*p* < 0.0001]).

From the mandatory reporting system, SARS-CoV-2 clinical data is also stratified geographically at the cantonal level. Therefore, we investigated the relationships between viral RNA loads from each location and the cases reported in the corresponding canton by computing Pearson correlation coefficients. We observed strong correlations with coefficients ranging from 0.55 to 0.93 (**Supplemental Table S4**).

### Lag times between clinical cases and viral RNA loads in wastewater

We used time-lagged cross-correlation to estimate lag times between wastewater viral loads and clinical cases for all four pathogens. Our analysis of weekly data showed that for RSV, the maximum value was reached at one week of lag, with wastewater data preceding clinical data (**Supplemental Fig. S5**). However, the computed coefficient was only slightly higher compared to the value obtained without lag (0.70 and 0.68, respectively) (**Supplemental Table S5**). In contrast, for IAV, IBV, and SARS-CoV-2, the highest Pearson correlation coefficient was identified with no lag.

To investigate if there was a lag within the first week, we used the clinical data obtained from the Swiss Federal Office of Public Health (FOPH) with daily resolution, which was provided for SARS-CoV-2, IAV, and IBV at the cantonal level. Therefore, we examined the correlations between viral RNA loads from each catchment and the reported cases in the corresponding canton. Our analysis did not reveal a consistent pattern of wastewater data preceding clinical data across all locations. Indeed, lag times for SARS-CoV-2 ranged from 0-5 days, for IAV from 0-7 days, and for IBV from 1-14 days, wastewater always being earlier.

Notably, we also observed lower correlation coefficients for SARS-CoV-2, when considering cantonal data with daily resolution instead of weekly aggregation. Coefficients ranged from 0.22-0.81 compared to 0.55-0.92. Moreover, as the 2023/2024 season was characterized by two waves of SARS-CoV-2, we also computed correlation coefficients for each wave separately, and we observed that in the second surge, values were generally ∼2.5 times lower, with coefficients ranging from 0.09-0.56.

## DISCUSSION

In this study, we report the successful development of a multiplex digital PCR (dPCR) assay targeting four common respiratory viruses – Influenza A (IAV) and B (IBV), Respiratory Syncytial Virus (RSV), SARS-CoV-2 (N1 and N2 genes) – and the internal control Murine Hepatitis Virus (MHV), an RNA virus used to monitor recovery efficiency. The six-plex dPCR assay enables absolute target quantification without the need for a standard curve for six targets simultaneously. A higher order of multiplexing can be achieved through additional channels or by amplitude-based multiplexing^14^, broadening the panel of detectable targets. Although we designed this assay for a 6-color system, it can be readily adapted to meet other requirements, by modifying fluorophores or changing channels. We applied our assay to Swiss wastewater samples within the scope of a previously established, national wastewater surveillance program^16^. We obtained four or five samples per week from 14 different wastewater treatment plants located across Switzerland, which collect the influent from more than one fourth of the Swiss residential population. Detecting viral RNA in wastewater is challenging due to low concentrations and the complexity of the matrix; indicating that efficient viral concentration methods are needed^17^. Here, we used MHV as internal control for extraction efficiency, and observed a mean recovery of 30%, which was close to the lower end of the range 26.7-65.7% found in a previous study that compared different concentration methods^18^. Notably, the extraction method used in our study which relies on direct capture, was not evaluated in the study. The reduced recovery could have been influenced by the contained wastewater volume employed (40 ml). Future efforts should focus on increasing sensitivity, by for example, increasing the volume of samples processed or improving recovery efficiency.

We detected and measured RNA from all assessed respiratory viruses, suggesting that our six-plex dPCR assay was specific to the selected genomic regions, and that our procedures were sufficiently sensitive to capture the clinically observed outbreaks for IAV, RSV, and SARS-CoV-2. The dPCR assay targeted both the SARS-CoV-2 N1 and N2 genes. Indeed, the use of various viral targets is generally advised, as the selection of only one could increase the rate of false-negatives^19^. However, this is not always feasible in practice, as it would greatly increase the required degree of multiplexing. Here, N1 and N2 markers showed strong agreement, despite N2 concentrations typically being higher than those of N1. Specifically, the N1 to N2 ratio was constantly below one until around January 2024, before returning to one. This observed discrepancy may have been triggered by mutations in the N1 probe binding region, which could reduce affinity, as proposed and demonstrated by Sun and colleagues^20^. However, we identified a point mutation (T28297C), positioned in the N1 forward primer binding site, which was present in 47.7% of the sequenced strains over the study period^21^. From July 2023 to January 2024, the percentage of strains carrying the mutation was 57.8%, which decreased to 4.3% from January to July 2024, corresponding to the timing of the N1/N2 ratio shift towards one. This observation suggests that T28297C lead to the decrease in N1/N2 ratio, highlighting how measured viral loads are impacted by the mutation landscape. By having multiple targets for a pathogen, decreasing primer/probe efficiency due to mutation can be more readily identified and primers or probes can be redesigned as required to ensure continuing assay performance.

We report that the 2023/2024 winter season in Switzerland was characterized by a substantial outbreak of SARS-CoV-2, with peak loads 10 to 100 times higher than those of other investigated respiratory viruses. This observed trend is consistent with previously reported loads^15,22^. The divergence in magnitude is potentially attributed to the higher transmissibility rate of SARS-CoV-2 compared to that of influenza and other seasonal respiratory viruses^23^. Notably, it could also be due to higher concentrations of SARS-CoV-2 RNA in feces of infected people relative to IAV, IBV, and RSV. Importantly, this winter season was dominated by the JN.1 variant^21^, which appears to be more transmissible than its parental virus^24,25^. In some catchments, we identified a second wave of SARS-CoV-2, which was smaller and driven by the KP.2 variant^26^.

Focusing on flu-season, IAV evidently prevailed over IBV, which was less abundant in wastewater and less reported in clinical settings. Despite the low number of positive cases of IBV, we detected its RNA in wastewater in 30% of the wastewater samples, which was not achievable by other studies^27,28^. Measurements of RSV in wastewater indicated the presence of a clear outbreak, which occurred between November 2023 and May 2024. Wastewater surveillance of RSV nicely demonstrates how wastewater data is valuable to get an insight into the epidemiological situation, in absence of a complete reporting system, as RSV infections are not subjected to mandatory declaration requirements in Switzerland.

For all viruses we observed occasional high measurement values. It is known that there are several sources of uncertainty in wastewater-based surveillance (WBS)^29^, and, despite attempts to control for some (e.g. inhibition), there are several which cannot be controlled for and their impact on concentration measurements remains to be elucidated. We therefore use a seven-day rolling median to determine a smoothed trend. Further work to understand specificities of wastewater viral concentration data could provide better methods for determining the average of technical replicates, generating a smooth signal or identifying outlier data points.

WBS has been extensively proposed as a useful tool for complementing clinical data, leading to a better understanding of respiratory diseases and identifying potential trends. In this study, we identified very strong correlations between wastewater viral loads and clinical cases, particularly for SARS-CoV-2 and IAV, as their occurrence was high during the monitoring period, and for RSV. The only exception was IBV, where we did not identify a strong correlation. In this case, wastewater data did not show a seasonal outbreak, whereas clinical cases collected through the mandatory reporting system indicated an outbreak (**Supplemental Fig. S3C**). Notably, cases from Sentinella and from the mandatory reporting system agreed only moderately. This might indicate that IBV was not causing a highly symptomatic infection or potentially had lower levels of viral shedding. Interestingly both the mandatory reporting and Sentinella systems indicate a roughly 80:20 ratio of IAV to IBV cases. The ability of the mandatory reporting system to capture the IBV outbreak is likely due to the greater number of cases reported in this way (n = 120’862 compared to n = 2’316).

Previous studies focusing on SARS-CoV-2 showed that the number of clinical cases follows viral titers in wastewater by variable lag times^30–33^. Our time-lagged cross-correlation analysis on four pathogens indicates that wastewater might serve as a leading indicator. However, we did not observe a uniform pattern of wastewater data consistently preceding clinical data across all sites, suggesting that some location-specific effects might exist. Differences might arise based on how representative the cantonal clinical data is when compared to the localized catchment viral load measurements. Moreover, several factors should be considered, such as population effects, including the proportion of vaccinated individuals and mobility behavior; and sewer network characteristics, like the influence of industrial discharge, and rainfall events^29^. Indeed, industrial wastewater might contain chemicals that could degrade RNA molecules; and rainfall events can introduce a dilution effect, leading to lower measured levels, which might fall below the limit of detection.

In the context of SARS-CoV-2, when comparing the relationship between wastewater loads and case data, we noticed that correlation coefficients were higher when clinical data were aggregated on a weekly level compared to daily data. This observation might be attributable to the intra-week variability in case reporting, or to a reduction of daily fluctuations of wastewater measurements when averaging over seven days. Additionally, as the monitoring period was characterized by two distinct waves, we observed a weaker relationship between wastewater loads and clinical cases during the spring-summer surge. This observation could be attributable to a decline in testing behavior during summer months or to the KP.2 variant potentially causing a less symptomatic infection.

Our study possesses some limitations. Despite reporting good assay sensitivity, considerable variations in pathogen concentrations in wastewater might lead to a reduction in sensitivity for detecting low-prevalence targets. Our results strongly suggest that WBS can be particularly valuable in settings where traditional surveillance systems are limited or inaccessible, such as resource-constrained areas or regions with inadequate healthcare facilities. However, we recognize that such an implementation and surveillance framework require a robust infrastructure, which may be challenging to establish in low- and middle-income countries.

Overall, we described a six-plex dPCR assay targeting four respiratory viruses and its application to Swiss wastewater. This allowed the identification and description of viral epidemiological trends at high temporal resolution over one entire year. The results demonstrated that tracking of priority clinical respiratory pathogens in wastewater provides data complementary to clinical-based surveillance. We believe that continued surveillance is crucial for long-term analysis and will provide insights into epidemiological trends and a more comprehensive understanding of pathogen dynamics over time, enabling more effective public health interventions.

## MATERIALS AND METHODS

### Experimental design

We developed a six-plex digital PCR (dPCR) assay targeting four respiratory viruses: SARS-CoV-2 N1 and N2 genes (N1 and N2), Influenza A M gene (IAV-M), Influenza B M gene (IBV-M), and Respiratory Syncytial Virus N gene (RSV-N), as well as Murine Hepatitis Virus M gene (MHV-M) which is used as an internal control for RNA extraction efficiency. The assay was applied to wastewater samples collected four or five times per week at 14 wastewater treatment plants from the 10^th^ of July 2023 until the 22^nd^ of July 2024.

### Wastewater treatment plants

Fourteen wastewater treatment plants (WWTPs) were monitored across Switzerland. The treatment plants provide service to approximately 2.3 million inhabitants across different regions (**Supplemental Table S1**), which corresponds to 26% of the national population.

### Wastewater sample processing and nucleic acid extraction

Raw 24-hour composite wastewater samples, routinely collected by WWTP personnel, were stored at 4°C and transported on ice to our laboratory at the Swiss Federal Institute of Aquatic Science and Technology (Eawag) in Dübendorf once a week. Samples were processed on the day of reception at Eawag which resulted in a delay between collection and processing of 2-7 days depending on the sample. Generally, five samples from each WWTP were processed weekly before 1st June 2024, then four per week afterwards.

For each sample, nucleic acids were extracted from 40 mL of wastewater using a modified version of the Wizard Enviro Total Nucleic Acid Kit (Promega Corporation, USA, Cat. No. A2991), as previously described^15^. A blank extraction control, consisting of tap water, was included in each extraction run to control for contamination during extraction. Nucleic acids were eluted in 80 µL RNase-free water and then purified using the OneStep PCR Inhibitor Removal Kit (Zymo Research, USA. Cat. No. D6030). Prior to dPCR analysis, extracts were further diluted in RNase-free water to minimize PCR inhibition (generally three-fold, however five-fold for Chur, Lugano, Basel and Geneva from August 2023 to March 2024). Typically, extracts were analyzed using dPCR immediately following their preparation on the same day. Extracts were stored for long-term preservation at −80°C.

### Positive controls

Positive controls used in dPCR assays were prepared by combining and mixing nucleic acid templates to achieve approximately 500 gene copies per µL for each target. Synthetic viral RNA was used for SARS-CoV-2, whereas gBlocks® were used for Influenza A and B, and RSV. Positive material for MHV was obtained by extracting RNA from cultured material (**Supplemental Table S6**).

### Digital PCR assays

All dPCR assays were developed on the Naica® system 6-color Crystal Digital PCR Prism-6 (Stilla Technologies, France). In this study, a six-plex assay, referred to as RESPV6, was developed. RESPV6 was created by merging two previous assays used in the Swiss monitoring program: i) a duplex assay (targeting the SARS-CoV-2 nucleoprotein gene locus 1 [N1] and the MHV matrix protein [M]), and ii) a previously described four-plex assay, known as RESPV4, with some modifications^15^. The RESPV4 was adapted for targeting the SARS-CoV-2 nucleoprotein gene locus 2 (N2), the Influenza A and B matrix protein genes (M), and the Respiratory Syncytial virus nucleoprotein gene (N). Primers and probes are listed and described in **Table 1** and were purchased from Microsynth AG or IDT (Switzerland).

**Table 1.**
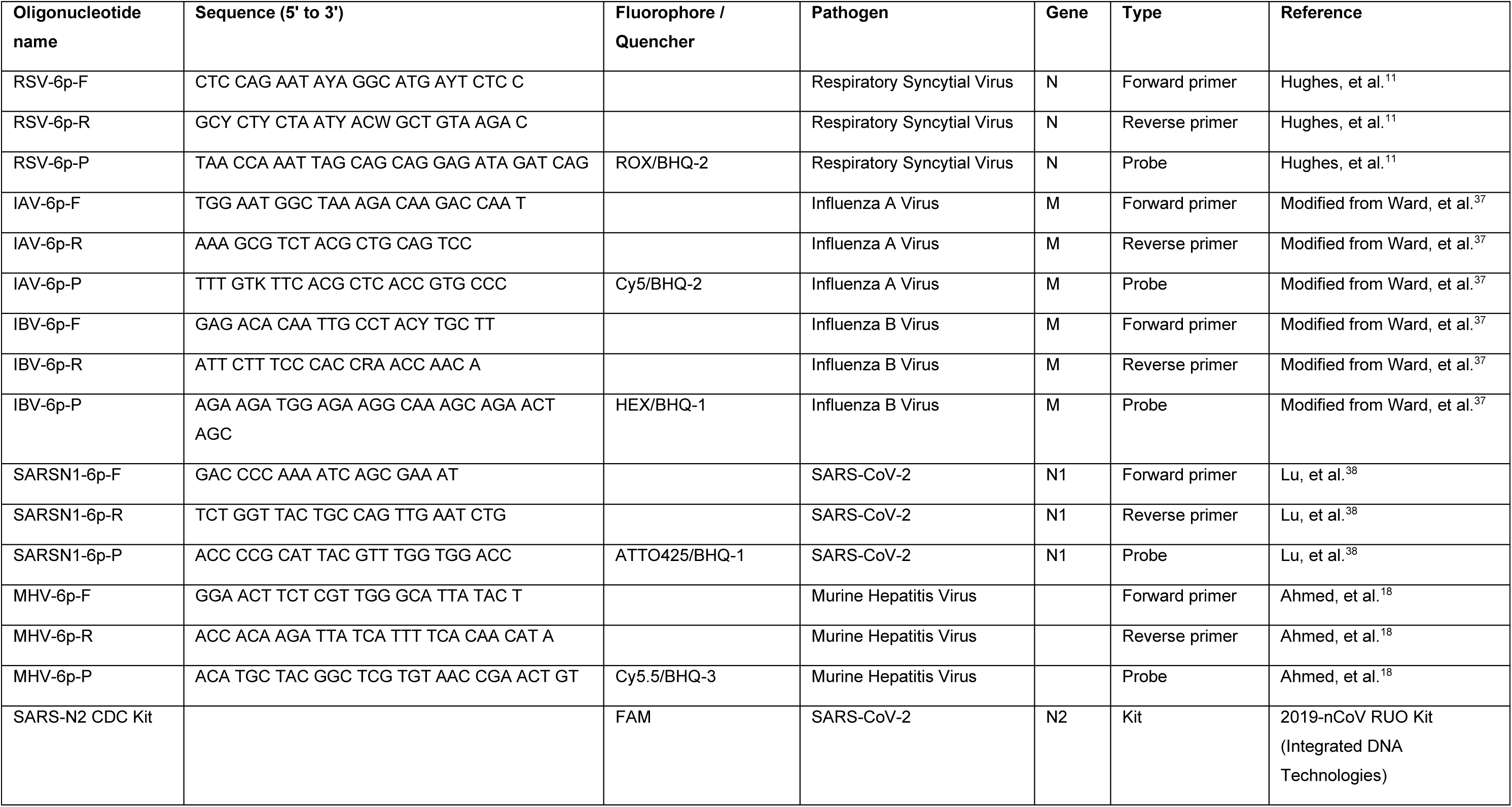
Primers and probes of the RESPV6 dPCR assay used in this study.

The RESPV6 assay was prepared using a total 27 µL pre-reaction volume, which consisted of 5.4 µL of template and 21.6 µL of mastermix. The mastermix was prepared as follows: qScript XLT One-Step RT-qPCR ToughMix (2x) (Quantabio, USA, Cat. No. 95132), 0.5 µM of each forward and reverse primer, 0.2 µM of each probe, SARS-N2 CDC Kit (0.125 µM of probe, 0.5 µM of primers, Integrated DNA Technologies, USA, Cat. No. 10006713,), 0.05 µM of fluorescein sodium salt (VWR, Cat. No. 0681-100G), and RNase-free water. The reaction volume (25 μL) was loaded into Sapphire chips (Stilla Technologies). Chips were loaded into a Geode (Stilla Technologies) which partitions the mastermix into droplets (12 min at 40°C; droplet volume: 0.519 nL) before thermocycling using the following conditions: reverse transcription (50°C for 1 h), enzyme activation (95°C for 5 min), and 40 cycles of denaturation (95°C for 30 s) and annealing/extension (57.5°C for 1 min).

One Geode can run three Sapphire chips in parallel, each containing four chambers, such that 12 samples can be analyzed at once. Generally, five wastewater nucleic acid extracts were run in technical duplicates, alongside a positive control and a no-template control (NTC). To pass quality control the positive control must have signal and the NTC must have fewer than three positive partitions. Additionally, there should be a minimum of 15’000 analyzable droplets. Reported data are the average of the technical replicates, although chamber failure occasionally occurred resulting in data from a singlet being taken. Reported data are the average of the technical replicates, although chamber failure occasionally occurred resulting in data from a singlet being taken. Our dPCR setup follows the digital MIQE (dMIQE) guidelines^34^ as detailed in **Annex 1**.

### PCR inhibition quality control

PCR inhibition in the RESPV6 assay was evaluated using a three-step approach as previously described^22^. First, the sample was analyzed to quantify the SARS-N1 target present in the wastewater matrix using the RESPV6 assay. In the second step, a known quantity of SARS-N1 RNA was spiked into the extract (approximately 750 gene copies per reaction), and the total SARS-N1 concentration was measured again with the same RESPV6 assay. Finally, inhibition was calculated using the following equation (1):

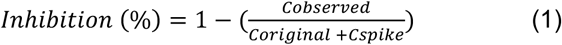

where: *C_observed_* is the concentration of SARS-N1 measured after spiking the sample with a known quantity of SARS-N1, *C_origina_*_l_ is the concentration of SARS-N1 in the unspiked samples, and *C_spike_* is the concentration of SARS-N1 added as the spike-in.

An inhibition value of 0% indicated no inhibition, suggesting that the PCR reaction efficiency was unaffected by the sample matrix. Conversely, a value of 100% indicated complete inhibition, where the PCR amplification of the measured concentration in the spiked assay resulted in no detection of the N1 assay in the wastewater extract. Values between 0 and 100% reflected partial inhibition, with higher values indicating higher levels of inhibition. Negative inhibition values, which are observed when C_observed_ is higher than the sum of C_original_ and C_spike_, are due to experimental variability, and are interpreted as an absence of inhibition. Our internal threshold for re-running a sample was set at 40%; samples with inhibition values above this threshold were considered inhibited and were subjected to re-analysis at a higher dilution to reduce inhibition depending on concentrations of SARS-CoV-2 in the wastewater. SARS-N1 inhibition served as a representative for the other targets. Measured RNA concentration was not corrected based on the quantitative estimates of inhibition.

### RNA extraction efficiency quality control

To determine the efficiency of RNA extraction, a known amount of viral control (Murine Hepatitis Virus strain MHV-A59, or MHV) was spiked into wastewater prior to processing (target concentration: 10’000 gene copies per ml of wastewater^8^) and measured using dPCR in the resultant extract. Culturing of MHV was performed in delayed brain tumor cells, at the École Polytechnique Fédérale de Lausanne (EPFL), as previously described^35^. Three out of every five samples (two out of every four samples after the 1^st^ June 2024) per week per WWTP were spiked with MHV. The recovery was determined by dividing the measured concentration by the estimated spiked concentration and expressing the result as a percentage. Measured RNA concentration was not corrected based on the quantitative estimates of the extraction efficiency.

### Data analysis and viral load calculation

All dPCR data were analyzed using the Crystal Miner Software version 4.0 (Stilla Technologies), which provides RNA quantities expressed as gene copies per microliter of reaction (gc/µl of reaction). Wastewater samples were defined as positive when the concentration was above the limit of detection (LoD), which was set at approximately five gc per reaction, and corresponds to a threshold of at least three positive partitions. Values were transformed to gene copies per liter of wastewater (gc/L_ww_), according to the following equation (2):

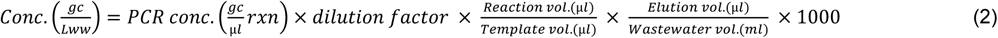

Viral RNA concentrations were adjusted by multiplying with the flow rate on the day of sampling at the influent of the WWTP and dividing by population in the catchment area. These values are referred to as viral loads and are expressed in gene copies per person per day (gc person^-1^ day^-1^).

A national median of viral loads was obtained by taking the median value over the WWTPs which have data for a particular day. The centered seven-day rolling median of these data points was then taken to visualize the trend.

### Comparison with clinical data from different sources

The Swiss Federal Office of Public Health (FOPH) collects clinical data about respiratory viruses mainly through two survey systems: the mandatory reporting system and the Swiss Sentinel System (Sentinella)^36^. The mandatory reporting system provides clinical data (i.e., clinical cases) for SARS-CoV-2, Influenza A and Influenza B. Publicly available data from the mandatory reporting system are grouped per week for the total Swiss population (8’855’062 inhabitants in 2024, including 39’677 inhabitants from Liechtenstein), and additionally the SARS-CoV-2 data is stratified by geographical location at the cantonal level. For this study, we additionally obtained data from the FOPH at daily resolution and stratified by canton. In contrast, the Sentinella system is a voluntary reporting system involving around 160 to 180 medical doctors and provides data for the four pathogens covered by our RESPV6 assay, including RSV^36^. Sentinella data are only available as total number of cases reported per week with no geographical location information available.

Since public case data for all the pathogens was only available at the national level, the national median of viral loads in wastewater was used for comparison. Additionally, due to the clinical data being grouped by week we computed a weekly average of our national wastewater load data by taking the mean daily value for each of the targets for each week. These national weekly averages were used for correlation analysis with case data.

Time-lagged cross-correlation analyses were performed to detect possible lag relationships between the viral loads in wastewater and clinical cases for each of the four pathogens. Wastewater data was shifted forward by a maximum of four weeks and cross-correlation was computed using Pearson correlation. Correlation analysis and time-lagged cross-correlation were also carried out on the daily resolution clinical data made available by the FOPH. Here, there was no requirement to aggregate wastewater viral load data. Case data at the cantonal level was matched to wastewater load data based on the canton where the WWTP is located (**Supplemental Table S1**).

## DATA AVAILABILITY

Digital PCR data are available for download from <u>wise.ethz.ch</u>. Only data which passed our described quality control processes are available here. All data is available on request. Weekly case data are available from the Federal Office of Public Health (FOPH) via a public <u>API</u>. Daily case data were specifically requested from the FOPH. All code used in the analysis is available on <u>GitHub</u>.

## Supporting information

Annex1

## ACKNOLEDGEMENTS

We would like to thank present and past members of the Laboratory Monitoring Team for developing the methods, processing samples and generating and analyzing data. Additionally, we thank present and past collaborators of the WISE (Wastewater-based Infectious disease Surveillance and Epidemiology) project for their support and valuable intellectual contributions, and NEXUS for their setting up and supporting the WISE database (wisedb.ethz.ch) which facilitated our analysis. We thank the technical staff of each WWTP for promptly sending us the wastewater samples on a weekly basis, and Anna Carratala Ripolles and Tamar Kohn for providing us with MHV.

## AUTHORS’ CONTRIBUTIONS

CO and TRJ conceptualized, supervised, and provided funding. MP and REM designed the study, performed analyses, prepared visualizations, and wrote the first draft of the manuscript. LC, JdK, CG, and SK contributed invaluable scientific insights. PS provided statistical support. AD, CH, AH, GL, LS, AW, and DY helped with sample processing, data generation and curation. All authors reviewed, edited, and approved the final version.

## FUNDING

This study was funded by the Swiss National Science Foundation (SNSF Sinergia Grant Nr. CRSII5_205933) and by the Swiss Federal Office of Public Health grant to CO and TRJ.

## CONFLICTS OF INTEREST

None.

## AUTHORS’ STATEMENT

We declare that all authors have seen and approved the manuscript and have contributed significantly to the work. Some data and figures from the manuscript presented here have been used to create a report to the primary funding body, the Swiss Federal Office of Public Health. The report was briefly made public (September – November 2024).

## Notes

### Competing Interest Statement

The authors have declared no competing interest.

### Author Declarations

Weekly case data are available from the Federal Office of Public Health (FOPH) via a public API (https://api.idd.bag.admin.ch/swagger-ui/api-doc.html). Daily case data are available on request to the FOPH and were used only for the purpose of correlation coefficient analyses. The datasets used in our study contain only aggregated data (no individual-level data have been used).

