## Supplemental Material for "A six-plex digital PCR assay for monitoring respiratory viruses in wastewater"

* Authors contributed equally

† Authors contributed equally

### Corresponding authors:

#### Annex 1. dMIQE Checklist

This file is available and submitted in Excel format.

#### TABLES

**Supplemental Table S1. Characteristics of selected locations and wastewater treatment plants in this study.** *Indicates treatment plants whose catchment area extends into one or more other cantons or other countries (i.e., STEP Aire into France).

| Location name | WWTP name | Population in catchment area | Canton | Cantonal population |
|---|---|---|---|---|
| Altenrhein | ARA Altenrhein* | 64000 | St. Gallen (SG) | 525967 |
| Basel | ARA Basel/Prorheno* | 268317 | Basel (BS) | 196786 |
| Bern | ARA Region Bern | 224557 | Bern (BE) | 1051437 |
| Chur | ARA Chur | 55000 | Graubünden (GR) | 202538 |
| Geneva | STEP Aire* | 454000 | Geneva (GE) | 514114 |
| Laupen | ARA Sensetal* | 62000 | Bern (BE) | 1051437 |
| Lausanne | STEP Vidy | 240000 | Vaud (VD) | 830431 |
| Lucern | ARA Buholz | 178555 | Lucerne (LU) | 424851 |
| Lugano | CDA Lugano | 124000 | Ticino (TI) | 354023 |
| Neuchatel | STEP Neuchatel | 40697 | Neuchâtel (NE) | 176571 |
| Porrentruy | STEP Porrentruy | 16500 | Jura (JU) | 73865 |
| Schwyz | ARA Schwyz | 31164 | Schwyz (SZ) | 164920 |
| Solothurn | ARA Zuchwil* | 95793 | Solothurn (SO) | 282408 |
| Zurich | ARA Werdhoelzli | 471000 | Zurich (ZH) | 1579967 |

**Supplemental Table S2.** **Comparison of PCR inhibition among 14 locations.** Pairwise comparisons were performed using the Dunn's test, Bonferroni correction was used to adjust for multiple testing. If the difference was statistically significant, asterisks were added in the last column (i.e., Significance) (***: p-value < 0.001, **: p-value < 0.01, and *: p-value < 0.05).

| **Comparison** | **Z** | **Unadjusted p-value** | **Adjusted p-value** | **Significance** |
|---|---|---|---|---|
| Altenrhein - Basel | -4.80E+00 | 1.58E-06 | 1.44E-04 | *** |
| Altenrhein - Bern | 8.93E-01 | 3.72E-01 | 1.00E+00 | |
| Basel - Bern | 5.61E+00 | 2.05E-08 | 1.86E-06 | *** |
| Altenrhein - Chur | -1.44E+00 | 1.50E-01 | 1.00E+00 | |
| Basel - Chur | 3.37E+00 | 7.41E-04 | 6.75E-02 | |
| Bern - Chur | -2.31E+00 | 2.10E-02 | 1.00E+00 | |
| Altenrhein - Geneva | -1.15E+00 | 2.52E-01 | 1.00E+00 | |
| Basel - Geneva | 3.70E+00 | 2.20E-04 | 2.00E-02 | * |
| Bern - Geneva | -2.02E+00 | 4.29E-02 | 1.00E+00 | |
| Chur - Geneva | 3.04E-01 | 7.61E-01 | 1.00E+00 | |
| Altenrhein - Laupen | -5.21E-01 | 6.02E-01 | 1.00E+00 | |
| Basel - Laupen | 4.29E+00 | 1.77E-05 | 1.61E-03 | ** |
| Bern - Laupen | -1.41E+00 | 1.60E-01 | 1.00E+00 | |
| Chur - Laupen | 9.22E-01 | 3.57E-01 | 1.00E+00 | |
| Geneva - Laupen | 6.24E-01 | 5.33E-01 | 1.00E+00 | |
| Altenrhein - Lausanne | -2.03E+00 | 4.24E-02 | 1.00E+00 | |
| Basel - Lausanne | 2.86E+00 | 4.21E-03 | 3.83E-01 | |
| Bern - Lausanne | -2.90E+00 | 3.72E-03 | 3.39E-01 | |
| Chur - Lausanne | -5.65E-01 | 5.72E-01 | 1.00E+00 | |
| Geneva - Lausanne | -8.78E-01 | 3.80E-01 | 1.00E+00 | |
| Laupen - Lausanne | -1.50E+00 | 1.33E-01 | 1.00E+00 | |
| Altenrhein - Lucerne | -5.43E-01 | 5.87E-01 | 1.00E+00 | |
| Basel - Lucerne | 4.28E+00 | 1.89E-05 | 1.72E-03 | ** |
| Bern - Lucerne | -1.43E+00 | 1.53E-01 | 1.00E+00 | |
| Chur - Lucerne | 9.03E-01 | 3.67E-01 | 1.00E+00 | |
| Geneva - Lucerne | 6.04E-01 | 5.46E-01 | 1.00E+00 | |
| Laupen - Lucerne | -2.09E-02 | 9.83E-01 | 1.00E+00 | |
| Lausanne - Lucerne | 1.48E+00 | 1.38E-01 | 1.00E+00 | |
| Altenrhein - Lugano | -5.73E-01 | 5.67E-01 | 1.00E+00 | |
| Basel - Lugano | 4.14E+00 | 3.44E-05 | 3.13E-03 | ** |
| Bern - Lugano | -1.44E+00 | 1.51E-01 | 1.00E+00 | |
| Chur - Lugano | 8.38E-01 | 4.02E-01 | 1.00E+00 | |
| Geneva - Lugano | 5.45E-01 | 5.86E-01 | 1.00E+00 | |
| Laupen - Lugano | -6.41E-02 | 9.49E-01 | 1.00E+00 | |
| Lausanne - Lugano | 1.40E+00 | 1.60E-01 | 1.00E+00 | |
| Lucerne - Lugano | -4.38E-02 | 9.65E-01 | 1.00E+00 | |
| Altenrhein - Neuchatel | 3.32E-01 | 7.40E-01 | 1.00E+00 | |
| Basel - Neuchatel | 5.13E+00 | 2.89E-07 | 2.63E-05 | *** |
| Bern - Neuchatel | -5.67E-01 | 5.71E-01 | 1.00E+00 | |
| Chur - Neuchatel | 1.77E+00 | 7.63E-02 | 1.00E+00 | |
| Geneva - Neuchatel | 1.48E+00 | 1.39E-01 | 1.00E+00 | |

| | | | | |
|---|---|---|---|---|
| Laupen - Neuchatel | 8.54E-01 | 3.93E-01 | 1.00E+00 | |
| Lausanne - Neuchatel | 2.37E+00 | 1.79E-02 | 1.00E+00 | |
| Lucerne - Neuchatel | 8.76E-01 | 3.81E-01 | 1.00E+00 | |
| Lugano - Neuchatel | 8.98E-01 | 3.69E-01 | 1.00E+00 | |
| Altenrhein - Porrentruy | -2.10E+00 | 3.59E-02 | 1.00E+00 | |
| Basel - Porrentruy | 2.74E+00 | 6.08E-03 | 5.53E-01 | |
| Bern - Porrentruy | -2.96E+00 | 3.08E-03 | 2.81E-01 | |
| Chur - Porrentruy | -6.51E-01 | 5.15E-01 | 1.00E+00 | |
| Geneva - Porrentruy | -9.61E-01 | 3.37E-01 | 1.00E+00 | |
| Laupen - Porrentruy | -1.58E+00 | 1.14E-01 | 1.00E+00 | |
| Lausanne - Porrentruy | -9.32E-02 | 9.26E-01 | 1.00E+00 | |
| Lucerne - Porrentruy | -1.56E+00 | 1.19E-01 | 1.00E+00 | |
| Lugano - Porrentruy | -1.48E+00 | 1.39E-01 | 1.00E+00 | |
| Neuchatel - Porrentruy | -2.43E+00 | 1.50E-02 | 1.00E+00 | |
| Altenrhein - Schwyz | -1.02E+00 | 3.06E-01 | 1.00E+00 | |
| Basel - Schwyz | 3.72E+00 | 2.03E-04 | 1.85E-02 | * |
| Bern - Schwyz | -1.88E+00 | 5.95E-02 | 1.00E+00 | |
| Chur - Schwyz | 3.93E-01 | 6.94E-01 | 1.00E+00 | |
| Geneva - Schwyz | 9.62E-02 | 9.23E-01 | 1.00E+00 | |
| Laupen - Schwyz | -5.14E-01 | 6.07E-01 | 1.00E+00 | |
| Lausanne - Schwyz | 9.56E-01 | 3.39E-01 | 1.00E+00 | |
| Lucerne - Schwyz | -4.94E-01 | 6.21E-01 | 1.00E+00 | |
| Lugano - Schwyz | -4.40E-01 | 6.60E-01 | 1.00E+00 | |
| Neuchatel - Schwyz | -1.35E+00 | 1.77E-01 | 1.00E+00 | |
| Porrentruy - Schwyz | 1.04E+00 | 3.00E-01 | 1.00E+00 | |
| Altenrhein - Solothurn | 5.26E+00 | 1.42E-07 | 1.29E-05 | *** |
| Basel - Solothurn | 9.66E+00 | 4.65E-22 | 4.23E-20 | *** |
| Bern - Solothurn | 4.35E+00 | 1.34E-05 | 1.22E-03 | ** |
| Chur - Solothurn | 6.59E+00 | 4.54E-11 | 4.13E-09 | *** |
| Geneva - Solothurn | 6.35E+00 | 2.17E-10 | 1.97E-08 | *** |
| Laupen - Solothurn | 5.75E+00 | 8.95E-09 | 8.15E-07 | *** |
| Lausanne - Solothurn | 7.21E+00 | 5.61E-13 | 5.10E-11 | *** |
| Lucerne - Solothurn | 5.78E+00 | 7.56E-09 | 6.88E-07 | *** |
| Lugano - Solothurn | 5.68E+00 | 1.34E-08 | 1.22E-06 | *** |
| Neuchatel - Solothurn | 4.96E+00 | 6.99E-07 | 6.36E-05 | *** |
| Porrentruy - Solothurn | 7.22E+00 | 5.31E-13 | 4.84E-11 | *** |
| Schwyz - Solothurn | 6.12E+00 | 9.32E-10 | 8.48E-08 | *** |
| Altenrhein - Zurich | -2.99E-01 | 7.65E-01 | 1.00E+00 | |
| Basel - Zurich | 4.51E+00 | 6.61E-06 | 6.01E-04 | *** |
| Bern - Zurich | -1.19E+00 | 2.35E-01 | 1.00E+00 | |
| Chur - Zurich | 1.14E+00 | 2.53E-01 | 1.00E+00 | |
| Geneva - Zurich | 8.46E-01 | 3.98E-01 | 1.00E+00 | |
| Laupen - Zurich | 2.22E-01 | 8.25E-01 | 1.00E+00 | |
| Lausanne - Zurich | 1.73E+00 | 8.43E-02 | 1.00E+00 | |
| Lucerne - Zurich | 2.43E-01 | 8.08E-01 | 1.00E+00 | |
| Lugano - Zurich | 2.81E-01 | 7.79E-01 | 1.00E+00 | |
| Neuchatel - Zurich | -6.31E-01 | 5.28E-01 | 1.00E+00 | |

| | | | | |
|---|---|---|---|---|
| Porrentruy - Zurich | 1.80E+00 | 7.20E-02 | 1.00E+00 | |
| Schwyz - Zurich | 7.31E-01 | 4.65E-01 | 1.00E+00 | |
| Solothurn - Zurich | -5.54E+00 | 3.05E-08 | 2.78E-06 | *** |

26

**Supplemental Table S3. Comparison of RNA extraction efficiency among 14 locations.** Pairwise comparisons were performed using the Dunn's test, Bonferroni correction was used to adjust for multiple testing. If the difference was statistically significant, asterisks were added in the last column (i.e., Significance) (***: p-value < 0.001, **: p-value < 0.01, and *: p-value < 0.05).

| Comparison | Z | Unadjusted p-value | Adjusted p-value | Significance |
|---|---|---|---|---|
| Altenrhein - Basel | 4.55E+00 | 5.31E-06 | 4.83E-04 | *** |
| Altenrhein - Bern | 2.85E+00 | 4.43E-03 | 4.03E-01 | |
| Basel - Bern | -1.73E+00 | 8.42E-02 | 1.00E+00 | |
| Altenrhein - Chur | 3.88E-01 | 6.98E-01 | 1.00E+00 | |
| Basel - Chur | -4.11E+00 | 3.88E-05 | 3.54E-03 | ** |
| Bern - Chur | -2.42E+00 | 1.54E-02 | 1.00E+00 | |
| Altenrhein - Geneva | 1.48E+00 | 1.38E-01 | 1.00E+00 | |
| Basel - Geneva | -3.04E+00 | 2.35E-03 | 2.14E-01 | |
| Bern - Geneva | -1.34E+00 | 1.81E-01 | 1.00E+00 | |
| Chur - Geneva | 1.08E+00 | 2.80E-01 | 1.00E+00 | |
| Altenrhein - Laupen | 3.22E+00 | 1.27E-03 | 1.16E-01 | |
| Basel - Laupen | -1.44E+00 | 1.49E-01 | 1.00E+00 | |
| Bern - Laupen | 3.19E-01 | 7.49E-01 | 1.00E+00 | |
| Chur - Laupen | 2.79E+00 | 5.32E-03 | 4.84E-01 | |
| Geneva - Laupen | 1.68E+00 | 9.28E-02 | 1.00E+00 | |
| Altenrhein - Lausanne | 1.87E+00 | 6.11E-02 | 1.00E+00 | |
| Basel - Lausanne | -2.70E+00 | 7.01E-03 | 6.38E-01 | |
| Bern - Lausanne | -9.75E-01 | 3.30E-01 | 1.00E+00 | |
| Chur - Lausanne | 1.46E+00 | 1.44E-01 | 1.00E+00 | |
| Geneva - Lausanne | 3.72E-01 | 7.10E-01 | 1.00E+00 | |
| Laupen - Lausanne | -1.31E+00 | 1.89E-01 | 1.00E+00 | |
| Altenrhein - Lucerne | 2.94E-01 | 7.69E-01 | 1.00E+00 | |
| Basel - Lucerne | -4.26E+00 | 2.02E-05 | 1.84E-03 | ** |
| Bern - Lucerne | -2.55E+00 | 1.07E-02 | 9.71E-01 | |
| Chur - Lucerne | -9.77E-02 | 9.22E-01 | 1.00E+00 | |
| Geneva - Lucerne | -1.19E+00 | 2.33E-01 | 1.00E+00 | |
| Laupen - Lucerne | -2.92E+00 | 3.45E-03 | 3.14E-01 | |
| Lausanne - Lucerne | -1.58E+00 | 1.14E-01 | 1.00E+00 | |
| Altenrhein - Lugano | -1.11E+00 | 2.66E-01 | 1.00E+00 | |
| Basel - Lugano | -5.60E+00 | 2.17E-08 | 1.97E-06 | *** |
| Bern - Lugano | -3.92E+00 | 8.85E-05 | 8.05E-03 | ** |
| Chur - Lugano | -1.48E+00 | 1.38E-01 | 1.00E+00 | |
| Geneva - Lugano | -2.57E+00 | 1.03E-02 | 9.34E-01 | |
| Laupen - Lugano | -4.31E+00 | 1.60E-05 | 1.46E-03 | ** |
| Lausanne - Lugano | -2.96E+00 | 3.07E-03 | 2.80E-01 | |
| Lucerne - Lugano | -1.40E+00 | 1.61E-01 | 1.00E+00 | |
| Altenrhein - Neuchatel | 9.92E-01 | 3.21E-01 | 1.00E+00 | |
| Basel - Neuchatel | -3.59E+00 | 3.32E-04 | 3.02E-02 | * |
| Bern - Neuchatel | -1.87E+00 | 6.18E-02 | 1.00E+00 | |
| Chur - Neuchatel | 5.90E-01 | 5.55E-01 | 1.00E+00 | |
| Geneva - Neuchatel | -5.08E-01 | 6.12E-01 | 1.00E+00 | |

| | | | | |
|---|---|---|---|---|
| Laupen - Neuchatel | -2.23E+00 | 2.60E-02 | 1.00E+00 | |
| Lausanne - Neuchatel | -8.89E-01 | 3.74E-01 | 1.00E+00 | |
| Lucerne - Neuchatel | 6.97E-01 | 4.86E-01 | 1.00E+00 | |
| Lugano - Neuchatel | 2.10E+00 | 3.61E-02 | 1.00E+00 | |
| Altenrhein - Porrentruy | 2.23E+00 | 2.56E-02 | 1.00E+00 | |
| Basel - Porrentruy | -2.30E+00 | 2.16E-02 | 1.00E+00 | |
| Bern - Porrentruy | -5.89E-01 | 5.56E-01 | 1.00E+00 | |
| Chur - Porrentruy | 1.82E+00 | 6.85E-02 | 1.00E+00 | |
| Geneva - Porrentruy | 7.43E-01 | 4.58E-01 | 1.00E+00 | |
| Laupen - Porrentruy | -9.17E-01 | 3.59E-01 | 1.00E+00 | |
| Lausanne - Porrentruy | 3.78E-01 | 7.06E-01 | 1.00E+00 | |
| Lucerne - Porrentruy | 1.94E+00 | 5.21E-02 | 1.00E+00 | |
| Lugano - Porrentruy | 3.31E+00 | 9.47E-04 | 8.61E-02 | |
| Neuchatel - Porrentruy | 1.26E+00 | 2.08E-01 | 1.00E+00 | |
| Altenrhein - Schwyz | 1.97E+00 | 4.85E-02 | 1.00E+00 | |
| Basel - Schwyz | -2.58E+00 | 9.91E-03 | 9.02E-01 | |
| Bern - Schwyz | -8.64E-01 | 3.88E-01 | 1.00E+00 | |
| Chur - Schwyz | 1.56E+00 | 1.18E-01 | 1.00E+00 | |
| Geneva - Schwyz | 4.77E-01 | 6.33E-01 | 1.00E+00 | |
| Laupen - Schwyz | -1.20E+00 | 2.31E-01 | 1.00E+00 | |
| Lausanne - Schwyz | 1.08E-01 | 9.14E-01 | 1.00E+00 | |
| Lucerne - Schwyz | 1.68E+00 | 9.28E-02 | 1.00E+00 | |
| Lugano - Schwyz | 3.05E+00 | 2.25E-03 | 2.05E-01 | |
| Neuchatel - Schwyz | 9.94E-01 | 3.20E-01 | 1.00E+00 | |
| Porrentruy - Schwyz | -2.69E-01 | 7.88E-01 | 1.00E+00 | |
| Altenrhein - Solothurn | 4.46E-01 | 6.56E-01 | 1.00E+00 | |
| Basel - Solothurn | -4.08E+00 | 4.48E-05 | 4.08E-03 | ** |
| Bern - Solothurn | -2.38E+00 | 1.72E-02 | 1.00E+00 | |
| Chur - Solothurn | 5.52E-02 | 9.56E-01 | 1.00E+00 | |
| Geneva - Solothurn | -1.03E+00 | 3.02E-01 | 1.00E+00 | |
| Laupen - Solothurn | -2.75E+00 | 6.03E-03 | 5.48E-01 | |
| Lausanne - Solothurn | -1.41E+00 | 1.57E-01 | 1.00E+00 | |
| Lucerne - Solothurn | 1.54E-01 | 8.77E-01 | 1.00E+00 | |
| Lugano - Solothurn | 1.54E+00 | 1.22E-01 | 1.00E+00 | |
| Neuchatel - Solothurn | -5.37E-01 | 5.91E-01 | 1.00E+00 | |
| Porrentruy - Solothurn | -1.78E+00 | 7.57E-02 | 1.00E+00 | |
| Schwyz - Solothurn | -1.52E+00 | 1.30E-01 | 1.00E+00 | |
| Altenrhein - Zurich | 3.85E-01 | 7.00E-01 | 1.00E+00 | |
| Basel - Zurich | -4.20E+00 | 2.63E-05 | 2.39E-03 | ** |
| Bern - Zurich | -2.48E+00 | 1.31E-02 | 1.00E+00 | |
| Chur - Zurich | -1.06E-02 | 9.92E-01 | 1.00E+00 | |
| Geneva - Zurich | -1.11E+00 | 2.66E-01 | 1.00E+00 | |
| Laupen - Zurich | -2.86E+00 | 4.30E-03 | 3.91E-01 | |
| Lausanne - Zurich | -1.50E+00 | 1.33E-01 | 1.00E+00 | |
| Lucerne - Zurich | 8.90E-02 | 9.29E-01 | 1.00E+00 | |
| Lugano - Zurich | 1.50E+00 | 1.33E-01 | 1.00E+00 | |
| Neuchatel - Zurich | -6.13E-01 | 5.40E-01 | 1.00E+00 | |

| | | | |
|---|---|---|---|
| Porrentruy - Zurich | -1.87E+00 | 6.18E-02 | 1.00E+00 |
| Schwyz - Zurich | -1.60E+00 | 1.09E-01 | 1.00E+00 |
| Solothurn - Zurich | -6.69E-02 | 9.47E-01 | 1.00E+00 |

Supplemental Table S4. **Correlation analysis between wastewater SARS-CoV-2 RNA loads from individual locations and clinical cases reported in the corresponding canton.** Correlation analysis was performed using Pearson's method.

| | | **Weekly case data** | | **Daily case data** | |
|---|---|---|---|---|---|
| **Location** | **Canton** | **Coefficient (r)** | **p-value** | **Coefficient (r)** | **p-value** |
| Altenrhein | SG | 0.86 | 9.43E-17 | 0.57 | 8.75E-24 |
| Basel | BS | 0.91 | 9.45E-22 | 0.79 | 2.05E-53 |
| Bern | BE | 0.89 | 1.99E-19 | 0.79 | 1.06E-55 |
| Chur | GR | 0.55 | 1.79E-05 | 0.22 | 6.48E-04 |
| Geneva | GE | 0.86 | 1.50E-16 | 0.76 | 8.53E-47 |
| Laupen | BE | 0.88 | 2.51E-18 | 0.71 | 1.08E-40 |
| Lausanne | VD | 0.57 | 8.60E-06 | 0.44 | 8.63E-13 |
| Lucerne | LU | 0.84 | 2.89E-15 | 0.65 | 9.38E-33 |
| Lugano | TI | 0.93 | 3.47E-24 | 0.81 | 1.48E-59 |
| Neuchatel | NE | 0.79 | 1.10E-12 | 0.59 | 7.28E-26 |
| Porrentruy | JU | 0.83 | 9.05E-15 | 0.54 | 1.64E-20 |
| Schwyz | SZ | 0.87 | 9.92E-18 | 0.61 | 1.24E-27 |
| Solothurn | SO | 0.82 | 1.23E-13 | 0.61 | 4.30E-27 |
| Zurich | ZH | 0.88 | 1.27E-18 | 0.66 | 2.02E-33 |

**Supplemental Table S5. Time-lagged cross-correlation analyses between viral RNA loads in wastewater and Sentinella clinical cases.** Correlation analysis was performed using Pearson's method. The highest correlation coefficient (r) for each pathogen and the corresponding lag time are visible in red.

| **Pathogen** | **Time lag (weeks)** | **Coefficient (r)** |
|---|---|---|
| Influenza A | 0 | 0.95 |
| Influenza A | 1 | 0.92 |
| Influenza A | 2 | 0.81 |
| Influenza A | 3 | 0.67 |
| Influenza A | 4 | 0.51 |
| Influenza B | 0 | 0.28 |
| Influenza B | 1 | 0.20 |
| Influenza B | 2 | 0.23 |
| Influenza B | 3 | 0.08 |
| Influenza B | 4 | 0.03 |
| Respiratory Syncytial Virus | 0 | 0.68 |
| Respiratory Syncytial Virus | 1 | 0.70 |
| Respiratory Syncytial Virus | 2 | 0.67 |
| Respiratory Syncytial Virus | 3 | 0.64 |
| Respiratory Syncytial Virus | 4 | 0.51 |
| SARS-CoV-2 | 0 | 0.87 |
| SARS-CoV-2 | 1 | 0.78 |
| SARS-CoV-2 | 2 | 0.69 |
| SARS-CoV-2 | 3 | 0.54 |
| SARS-CoV-2 | 4 | 0.38 |

**Supplemental Table S6. Characteristics of nucleic acids used as positive material in this study.**

| Viral target | Targeted sequence | Source |
|---|---|---|
| SARS-CoV-2 | Whole genome, GISAID: EPI_ISL_7718520 | Control 51 (B.1.1.529/BA.2) (Twist Bioscience) |
| MHV-A59 | Whole genome | RNA extracted from cultured viral material (provided by Dr. Anna Carratala Ripolles, EPFL) |
| Influenza A | tatgttctctctatcgttccgtcaggccccctcaaagccgagatagcgcagagacttgaagatgtttttgcagggaaaaacaccgatcttgaggcactcatgg aatggctaaagacaagaccaatcctgtcacctctgactaaggggattttaggatttgtgttcacgctcaccgtgcccagtgagcgaggactgcagcgtaga cgctttgtccagaatgccctcaatgggaatggtgacccgaacaacatggacaaagcggtcaaactgtacagaaaacttaaaagggaaataacattccac ggggcca | gBlock (Integrated DNA Technologies, IDT) |
| Influenza B | atgtcgctgtttggagacacaattgcctacctgctttcactaatagaagatggagaaggcaaagcagaactagctgaaaaattacactgttggttcggtggg aaagaatttgacctagattctgctttggaatggataaaaaacaaaaggtgcctaactgatatacaaaaagcactaattggtgcctctatatgctttttaaaacc caa | gBlock (IDT) |
| Respiratory Syncytial Virus | gtattaacattatcaagcttgacatcagaaatacaagtcaatattgagatagaatctagaaagtcctacaaaaaaatgctaaaagagatgggagaagtgg ctccagaatataggcatgattctccagactgtgggatgataatactgtgtatagctgcccttgtaataaccaaattagcagcaggagatagatcaggtcttac agcagtaattaggagggcaaacaatgtcttaaaaaacgaaataaaacgctacaagggcctaataccaaaagacatagccaacagtttttatgaagtattt gaaaaataccctcatctt | gBlock (IDT) |

**Supplemental Table S7. Values excluded from graphical representations.** This table includes all values that are not visible in the graphical representation. NA: not applicable.

| Figure | Location | Target | Date of collection | Value |
|---|---|---|---|---|
| Fig. 3 | Altenrhein | SARS-N1 | 14-Jan-24 | 1.50E+09 |
| Fig. 3 | Altenrhein | SARS-N2 | 14-Jan-24 | 1.59E+09 |
| Fig. 3 | Lucerne | SARS-N1 | 09-Dec-23 | 1.33E+09 |
| Fig. 3 | Lucerne | SARS-N1 | 11-Dec-23 | 1.66E+09 |
| Fig. 3 | Lucerne | SARS-N2 | 06-Dec-23 | 9.97E+08 |
| Fig. 3 | Lucerne | SARS-N2 | 09-Dec-23 | 1.74E+09 |
| Fig. 3 | Lucerne | SARS-N2 | 11-Dec-23 | 2.12E+09 |
| Fig. 3 | Chur | SARS-N1 | 22-Oct-23 | 7.39E+09 |
| Fig. 3 | Chur | SARS-N2 | 22-Oct-23 | 1.21E+10 |
| Fig. 3 | Schwyz | SARS-N1 | 15-Nov-23 | 1.46E+09 |
| Fig. 3 | Schwyz | SARS-N1 | 05-Dec-23 | 9.40E+08 |
| Fig. 3 | Schwyz | SARS-N2 | 15-Nov-23 | 1.76E+09 |
| Fig. 3 | Schwyz | SARS-N2 | 20-Nov-23 | 9.11E+08 |
| Fig. 3 | Schwyz | SARS-N2 | 05-Dec-23 | 1.09E+09 |
| Fig. 3 | Zurich | SARS-N1 | 12-Nov-23 | 1.59E+09 |
| Fig. 3 | Zurich | SARS-N2 | 12-Nov-23 | 1.90E+09 |
| Fig. 3 | Zurich | SARS-N2 | 03-Dec-23 | 9.17E+08 |
| Fig. 3 | Zurich | SARS-N2 | 23-Dec-23 | 9.24E+08 |
| Fig. 3 | Solothurn | SARS-N2 | 07-Dec-23 | 9.91E+08 |
| Fig. 3 | Lugano | SARS-N1 | 11-Nov-23 | 1.21E+09 |
| Fig. 3 | Lugano | SARS-N2 | 11-Nov-23 | 1.32E+09 |
| Fig. 3 | Lugano | SARS-N2 | 20-Nov-23 | 9.41E+08 |
| Fig. 3 | Lugano | SARS-N2 | 25-Nov-23 | 9.06E+08 |
| Fig. 3 | Lugano | SARS-N2 | 30-Nov-23 | 9.16E+08 |
| Fig. 3 | Lugano | SARS-N2 | 06-Dec-23 | 9.47E+08 |
| Fig. 3 | Lugano | SARS-N2 | 14-Dec-23 | 9.76E+08 |
| Fig. 3 | Geneva | SARS-N1 | 12-Dec-23 | 9.19E+08 |
| Fig. 3 | Geneva | SARS-N2 | 12-Dec-23 | 1.08E+09 |
| Fig. 3 | Neuchatel | SARS-N1 | 15-Jun-24 | 9.28E+08 |
| Fig. 3 | Neuchatel | SARS-N2 | 15-Jun-24 | 9.54E+08 |
| Fig. 3 | Lausanne | SARS-N1 | 23-Jun-24 | 2.33E+09 |
| Fig. 3 | Lausanne | SARS-N2 | 26-Nov-23 | 1.08E+09 |
| Fig. 3 | Lausanne | SARS-N2 | 11-Dec-23 | 1.02E+09 |
| Fig. 3 | Lausanne | SARS-N2 | 23-Jun-24 | 2.46E+09 |
| Fig. 4 | Altenrhein | IAV-M | 14-Jan-24 | 1.58E+08 |
| Fig. 4 | Altenrhein | IAV-M | 29-Jan-24 | 1.71E+08 |
| Fig. 4 | Altenrhein | RSV-N | 14-Jan-24 | 1.58E+08 |
| Fig. 4 | Altenrhein | RSV-N | 29-Jan-24 | 1.60E+08 |
| Fig. 4 | Bern | IAV-M | 26-Jul-23 | 2.17E+08 |
| Fig. 4 | Schwyz | IAV-M | 22-Jan-24 | 1.01E+08 |
| Fig. 4 | Schwyz | IAV-M | 30-Jan-24 | 6.08E+07 |
| Fig. 4 | Zurich | RSV-N | 26-Nov-23 | 1.08E+08 |

| | | | | |
|---|---|---|---|---|
| Fig. 4 | Solothurn | IBV-M | 13-Jan-24 | 6.49E+07 |
| Fig. 4 | Solothurn | IBV-M | 13-Feb-24 | 7.57E+07 |
| Fig. 4 | Solothurn | IBV-M | 15-Apr-24 | 1.22E+08 |
| Fig. 4 | Solothurn | IBV-M | 26-Jun-24 | 7.64E+07 |
| Fig. 4 | Solothurn | RSV-N | 22-Jul-23 | 7.59E+08 |
| Fig. 4 | Solothurn | RSV-N | 24-Jul-23 | 3.59E+08 |
| Fig. 4 | Lugano | IAV-M | 24-Dec-23 | 1.05E+08 |
| Fig. 4 | Lugano | IAV-M | 25-Dec-23 | 7.32E+07 |
| Fig. 4 | Lugano | IAV-M | 31-Dec-23 | 6.02E+07 |
| Fig. 4 | Neuchatel | IAV-M | 21-Jan-24 | 8.18E+07 |
| Fig. 4 | Porrentruy | IAV-M | 19-Jul-23 | 2.29E+08 |
| Fig. 4 | Porrentruy | IAV-M | 24-Jul-23 | 6.06E+08 |
| Fig. 4 | Porrentruy | IBV-M | 24-Jul-23 | 3.09E+08 |
| Fig. 5 and Fig. S3 | NA | IAV | 24-Jan-24 | 3.30E+07 |
| Fig. 5 | NA | RSV | 24-Dec-23 | 1.04E+07 |
| Fig. 5 | NA | RSV | 31-Dec-23 | 1.02E+07 |
| Fig. 5 | NA | RSV | 2-Jan-24 | 1.43E+07 |
| Fig. 5 | NA | RSV | 4-Jan-24 | 1.14E+07 |
| Fig. 5 | NA | RSV | 8-Jan-24 | 1.07E+07 |
| Fig. 5 | NA | RSV | 21-Jan-24 | 1.05E+07 |
| Fig. 5 | NA | RSV | 29-Jan-24 | 1.14E+07 |
| Fig. 5 and Fig. S3 | NA | SARS | 25-Nov-23 | 4.40E+08 |
| Fig. 5 and Fig. S3 | NA | SARS | 26-Nov-23 | 4.38E+08 |
| Fig. 5 and Fig. S3 | NA | SARS | 27-Nov-23 | 4.97E+08 |
| Fig. 5 and Fig. S3 | NA | SARS | 4-Dec-23 | 4.61E+08 |
| Fig. 5 and Fig. S3 | NA | SARS | 7-Dec-23 | 4.27E+08 |
| Fig. S1 | Altenrhein | NA | 30-Aug-23 | 2.55 |
| Fig. S1 | Altenrhein | NA | 1-Sep-23 | 2.28 |
| Fig. S1 | Altenrhein | NA | 19-Mar-24 | 6.04 |
| Fig. S1 | Basel | NA | 3-Sep-23 | 2.75 |
| Fig. S1 | Lucerne | NA | 1-Sep-23 | 2.07 |
| Fig. S1 | Lucerne | NA | 4-Sep-23 | 2.25 |
| Fig. S1 | Bern | NA | 25-Jul-23 | 2.24 |
| Fig. S1 | Bern | NA | 28-Mar-24 | 2.08 |
| Fig. S1 | Schwyz | NA | 11-Mar-24 | 2.50 |
| Fig. S1 | Schwyz | NA | 17-Mar-24 | 3.50 |
| Fig. S1 | Laupen | NA | 1-Sep-23 | 2.11 |
| Fig. S1 | Laupen | NA | 3-Sep-23 | 2.02 |
| Fig. S1 | Laupen | NA | 4-Sep-23 | 2.23 |
| Fig. S1 | Laupen | NA | 26-Feb-24 | 2.25 |
| Fig. S1 | Laupen | NA | 5-Mar-24 | 3.94 |
| Fig. S1 | Laupen | NA | 25-Mar-24 | 2.10 |
| Fig. S1 | Laupen | NA | 2-Apr-24 | 3.49 |
| Fig. S1 | Laupen | NA | 16-May-24 | 2.00 |
| Fig. S1 | Solothurn | NA | 1-Mar-24 | 4.06 |
| Fig. S1 | Solothurn | NA | 8-Mar-24 | 2.20 |

| | | | | |
|---|---|---|---|---|
| Fig. S1 | Solothurn | NA | 18-Mar-24 | 2.00 |
| Fig. S1 | Solothurn | NA | 7-Apr-24 | 2.58 |
| Fig. S1 | Solothurn | NA | 11-Apr-24 | 2.76 |
| Fig. S1 | Solothurn | NA | 13-Apr-24 | 2.80 |
| Fig. S1 | Solothurn | NA | 25-Apr-24 | 2.00 |
| Fig. S1 | Lugano | NA | 30-Aug-23 | 2.01 |
| Fig. S1 | Lugano | NA | 27-Feb-24 | 3.00 |
| Fig. S1 | Lugano | NA | 1-Mar-24 | 6.10 |
| Fig. S1 | Lugano | NA | 18-Mar-24 | 2.00 |
| Fig. S1 | Lugano | NA | 1-Apr-24 | 2.05 |
| Fig. S1 | Porrentruy | NA | 18-Feb-24 | 2.73 |
| Fig. S1 | Porrentruy | NA | 20-Feb-24 | 4.03 |
| Fig. S1 | Porrentruy | NA | 5-Mar-24 | 2.41 |
| Fig. S1 | Porrentruy | NA | 17-Mar-24 | 3.00 |
| Fig. S1 | Porrentruy | NA | 31-Mar-24 | 2.00 |
| Fig. S1 | Porrentruy | NA | 3-Apr-24 | 2.00 |
| Fig. S1 | Porrentruy | NA | 5-Apr-24 | 2.00 |
| Fig. S1 | Porrentruy | NA | 13-Apr-24 | 5.13 |
| Fig. S1 | Porrentruy | NA | 17-Apr-24 | 4.58 |
| Fig. S1 | Porrentruy | NA | 19-Apr-24 | 2.62 |
| Fig. S1 | Porrentruy | NA | 13-May-24 | 2.00 |

45

#### FIGURES

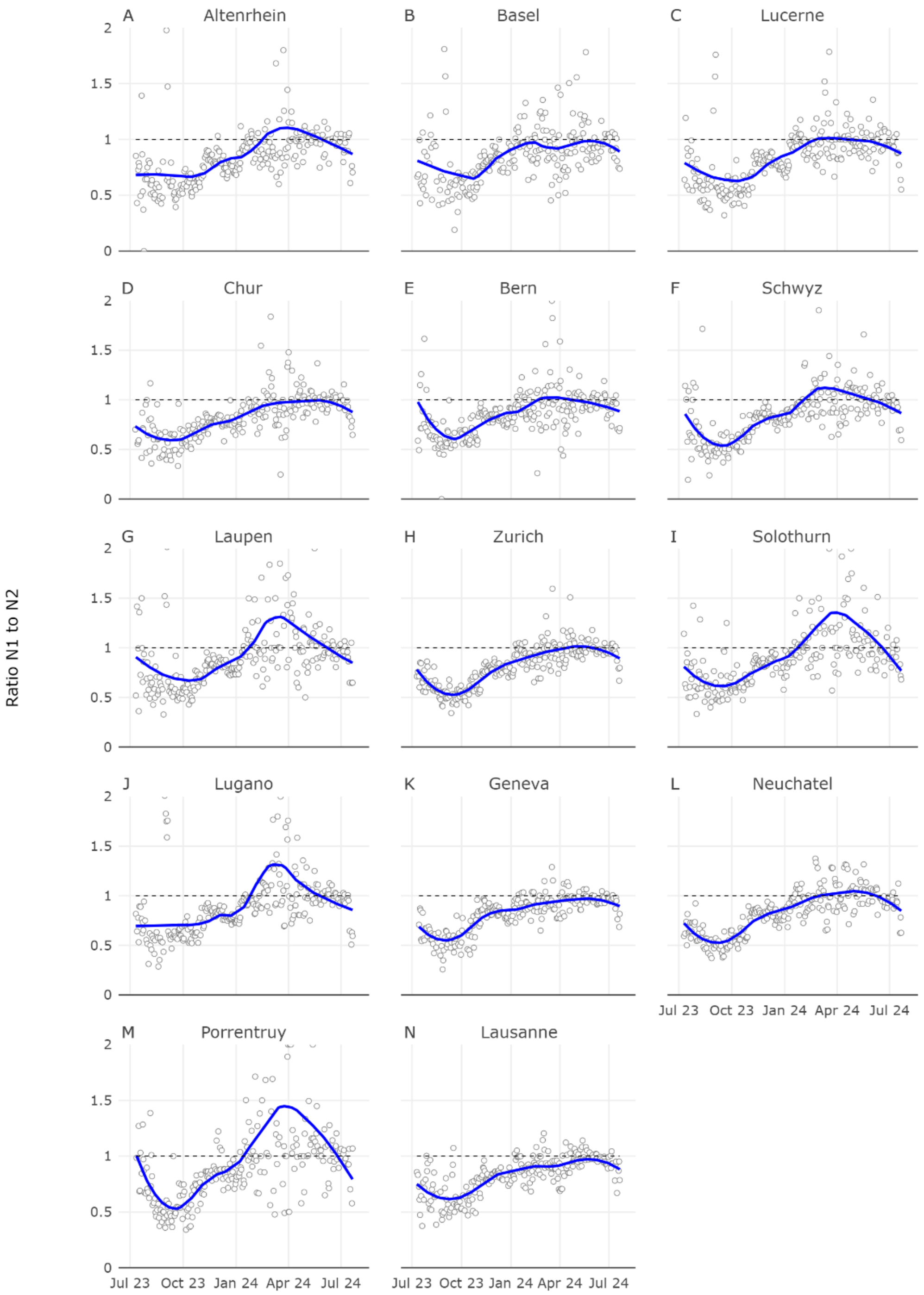

**Supplemental Figure S1. Variation of N1 to N2 ratio over time.** Grey circles indicate the N1 to N2 ratio. LOESS was used for fitting the smooth blue line. The dashed grey line indicates a ratio of one. Some high ratios are beyond the plotted y-max values. They are listed in **Supplemental Table S7**.

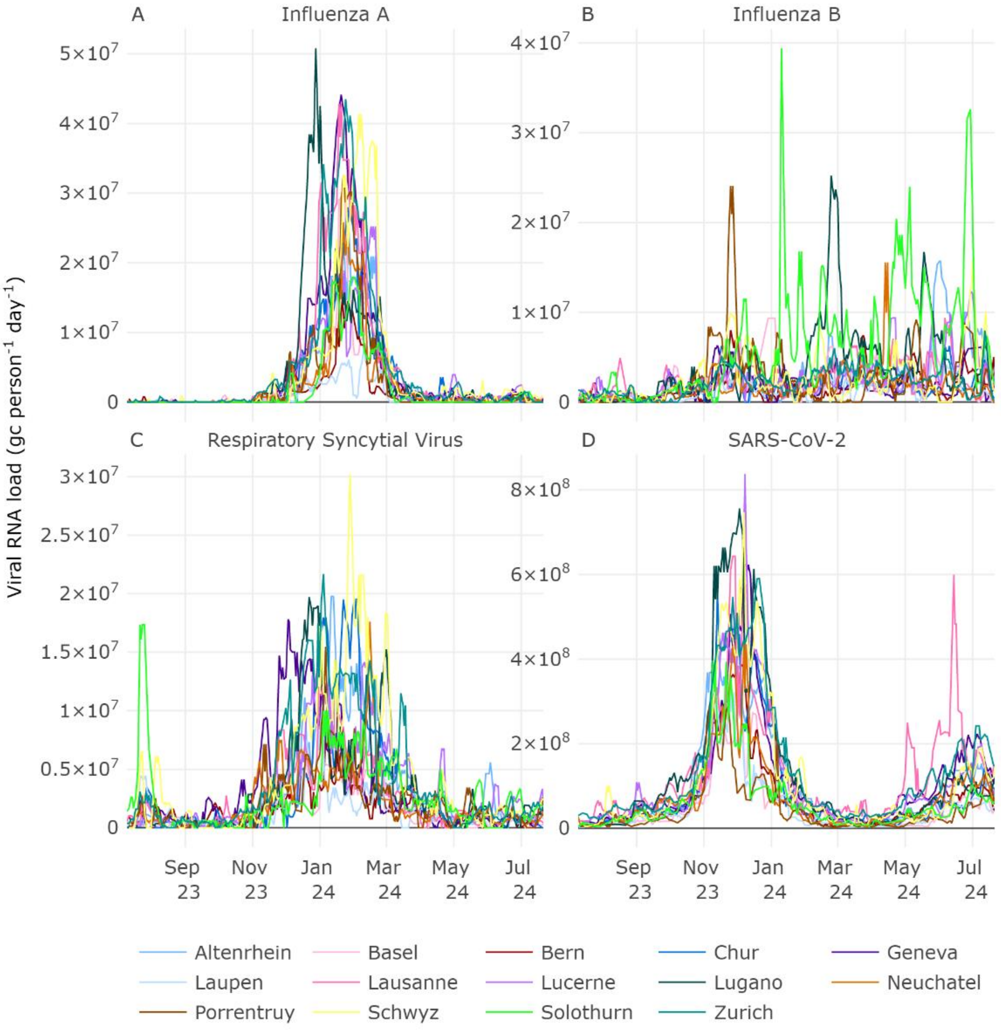

**Supplemental Figure S2. Comparison of Influenza A, Influenza B, RSV and SARS-CoV-2 RNA loads in wastewater across 14 Swiss locations.** Viral RNA loads are in gene copies per person per day and shown on the y-axis. The 7-day rolling median trend is plotted.

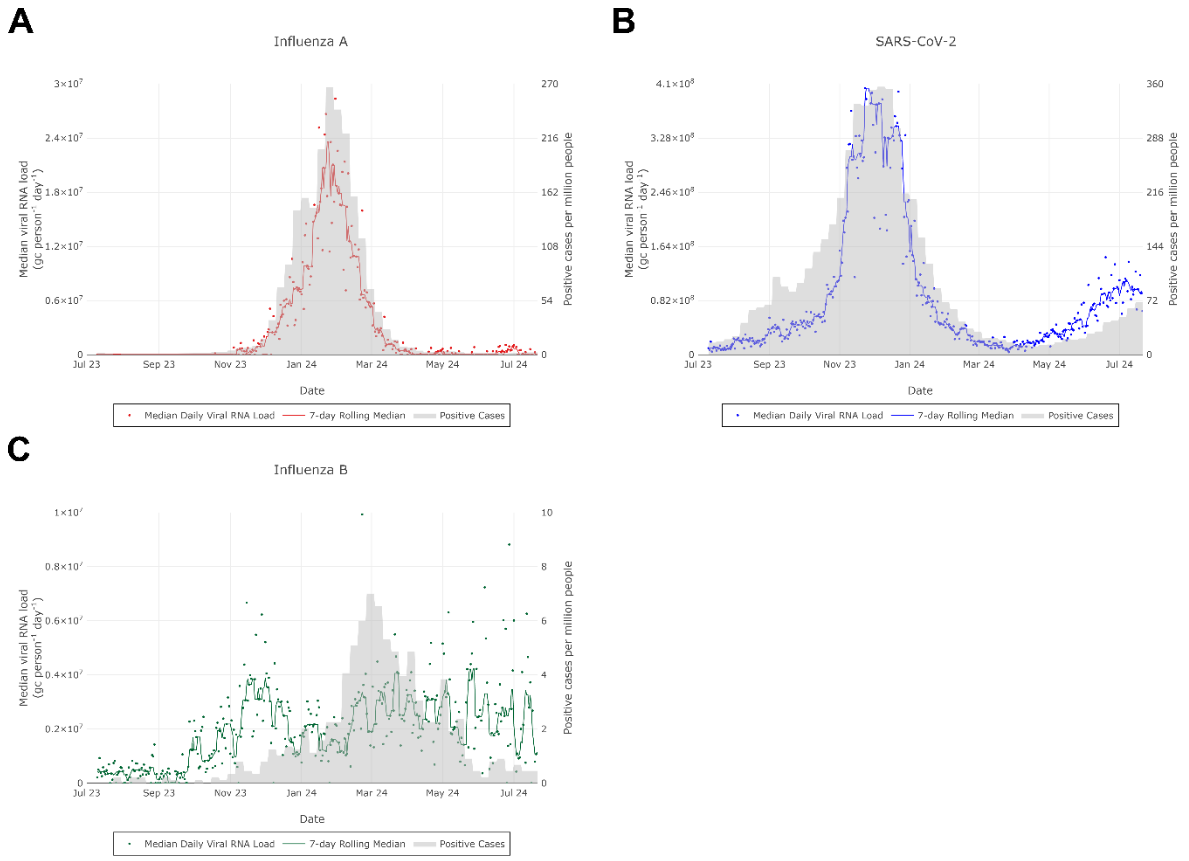

**Supplemental Figure S3. Comparison of viral RNA loads in wastewater and number of cases from mandatory reporting system.** Positive cases, expressed in positive cases per million people, refer to the entire Swiss territory, and were taken from the mandatory reporting database. Cases are shown using light grey bars. Some high load values are beyond the plotted y-max values. They are listed in **Supplemental Table S7**. This figure has been formatted using Inkscape.

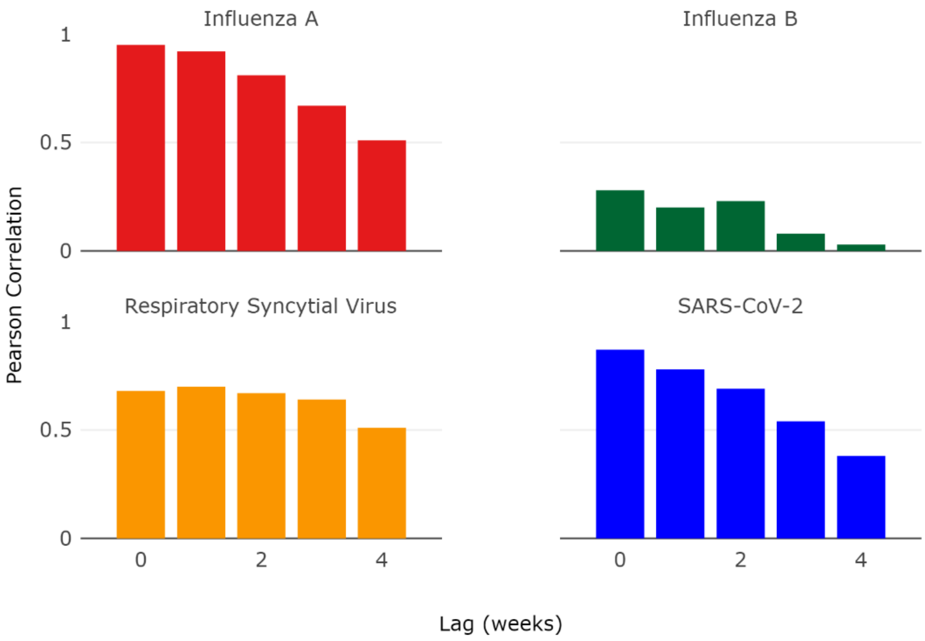

**Supplemental Figure S4. Time-lagged Pearson correlation coefficients between viral RNA loads in wastewater and Sentinella clinical cases.**
